# Performance of Large Language Models in Automated Medical Literature Screening: A Systematic Review and Meta-analysis

**DOI:** 10.64898/2026.03.17.26348656

**Authors:** Chenggong Xie, Weichang Kong, Liuting Pi, Diaoxin Qi, Yuxuan Yang, Bin Wang, Hao Liang

**Author notes:** **Corresponding author:** Hao Liang; Bin Wang.

## Abstract

**Objective:** To systematically evaluate the diagnostic performance of large language models (LLMs) in automated medical literature screening and to determine their potential role in supporting evidence synthesis workflows.

**Methods:** A systematic review and meta-analysis was conducted according to PRISMA DTA guidance. PubMed, Web of Science, Embase, the Cochrane Library and Google Scholar were searched from 1 January 2022 to 17 November 2025. Studies assessing LLMs for automated title and abstract screening or full-text eligibility assessment in medical literature were included. Diagnostic accuracy metrics were extracted and pooled using a bivariate random effects model and hierarchical summary receiver operating characteristic (HSROC) analysis. Subgroup analyses and meta-regression were performed to explore sources of heterogeneity.

**Results:** Eighteen studies published between 2023 and 2025 were included. In title and abstract screening, the pooled sensitivity was 0.92 and pooled specificity was 0.94. The SROC area under the curve (AUC) reached 0.98. In full-text screening, pooled sensitivity and specificity both reached 0.99 and the AUC was 0.99. Prompt strategies incorporating examples or chain-of-thought reasoning significantly improved sensitivity. Across studies, most models were deployed without task specific fine tuning and still achieved strong performance. Subgroup analyses and meta regression did not identify significant sources of heterogeneity. Many studies also reported substantial efficiency gains, including large reductions in screening workload, time and cost.

**Conclusion:** LLMs demonstrate high diagnostic accuracy for automated medical literature screening, particularly in full-text assessment. These models show strong potential as high sensitivity assistive tools that can substantially reduce manual screening burden while supporting evidence synthesis. Further methodological optimization and validation in large scale real-world settings are required to establish their long term role in evidence-based medicine.

## 1. Introduction

Medical literature serves as the primary repository of medical knowledge and scientific insight and plays a central role in advancing healthcare and biomedical research(1). The retrieval and screening of relevant medical literature constitute a fundamental step in evidence-based medicine and biomedical knowledge management(2). In most cases, researchers rely on manual processes to search, screen, and curate relevant literature sets(3). However, as the volume of biomedical publications continues to expand at an unprecedented pace, manual screening of medical literature has become increasingly impractical and unsustainable(4). With advances in artificial intelligence (AI), automated approaches to medical literature screening have gradually been introduced into real world research workflows(5).

To improve efficiency, a range of automated methods has been developed to support medical literature screening, including rule-based filtering, traditional machine learning (TML) classifiers, and more recently, deep learning (DL) models(6-8). Although these approaches have shown promise in reducing screening workload, they typically depend on large amounts of task specific labeled data and demonstrate limited adaptability across different clinical topics and study designs(9, 10). Moreover, many existing models struggle to capture the nuanced semantic and methodological criteria that underpin eligibility decisions in medical literature screening.

Over the past three years, the rapid development and deployment of large language models (LLMs) have introduced new opportunities for automated medical literature screening(11). Pretrained on large scale textual corpora and equipped with instruction following and contextual reasoning capabilities, LLMs can interpret complex eligibility criteria and apply them directly to medical literature in zero-shot or few-shot settings(12). Unlike conventional classifiers, these models function as general purpose text understanding systems, enabling flexible adaptation to diverse screening tasks without the need for extensive retraining(13).

Despite growing interest in this area, empirical evidence regarding the performance of LLMs in automated medical literature screening remains fragmented(14). Existing studies differ substantially in screening tasks, medical domains, evaluation metrics, and implementation strategies(15). Most investigations are confined to single datasets or case studies, and no systematic review and meta-analysis has comprehensively synthesized the available evidence to quantify model performance or to explore sources of heterogeneity. Rigorous evidence synthesis is therefore required to guide the appropriate and responsible application of LLMs in medical literature screening.

Accordingly, this study conducts a systematic review and meta-analysis of LLM-based tools for automated medical literature screening, with the aim of determining whether these models can meaningfully complement or partially replace manual screening while improving efficiency and reducing human workload.

## 2. Methods

This systematic review was conducted in accordance with the PRISMA-DTA statement(16). As the analyses were based exclusively on aggregated data extracted from previously published non-clinical or laboratory research studies, ethical approval and informed consent from individual participants were not necessary.

### 2.1 Data Sources

We developed database specific search strategies for five literature sources, including PubMed, Web of Science, Embase, the Cochrane Library, and Google Scholar, covering the period from 1 January 2022 to 17 November 2025. The start date was chosen because LLMs began to emerge as a major research focus in 2022 and were increasingly applied across the full research workflow, including literature screening and other scientific data processing tasks. The search combined controlled vocabulary terms from MeSH and Emtree with free text terms. Relevant concepts, along with their synonyms and related terms, were used to capture studies referring to LLMs and literature screening. Full details of the search strategies are provided in Supplementary Material 1.

### 2.2 Inclusion and exclusion criteria

Studies were selected according to predefined eligibility criteria. Eligible studies were required to meet all of the following inclusion criteria: 1) the study evaluated the use of LLMs for automated or semi-automated medical literature screening, including title/abstract screening and/or full-text eligibility assessment; 2) the screening task was conducted within a medical, biomedical, or health-related research context, such as systematic reviews; 3) general-purpose LLMs were applied in prompt-based settings, with or without minimal task-specific adaptation; 4) the study reported quantitative performance metrics relevant to screening accuracy or efficiency, including but not limited to sensitivity, specificity, precision, F1-score and accuracy and 5) the study was an original peer-reviewed journal article or conference proceeding in English.

Studies were excluded if they met any of the following exclusion criteria: 1) the publication was not an original research article (e.g., review, editorial, commentary, protocol, or perspective); 2) the study did not provide sufficient quantitative information to extract or calculate performance metrics for literature screening; 3) duplicate reports describing the same dataset or experiment or 4) the full-text was unavailable after reasonable efforts to obtain it.

### 2.3 Study selection and data extraction

Two reviewers, WCK and LTP, independently assessed the full texts of all eligible studies and extracted the data. In cases of disagreement, discussions were led by the principal investigator, CGX, until consensus was reached. For each included study, the following information was collected: first author and year of publication, type of included studies, number of included studies, search content, method used to establish the gold standard, number of articles required to be screened, and number of articles ultimately included.

For the LLMs component of each study, additional details were extracted, including the models used and their versions, deployment approach, prompt type, whether model fine tuning was performed, alignment methods as well as performance and efficiency metrics.

### 2.4 Quality assessment

To evaluate the methodological quality and risk of bias of the included models and studies, we applied the Prediction Model Risk of Bias Assessment Tool + AI tool (PROBAST + AI) and the Quality Assessment of Diagnostic Accuracy Studies-2 tool, respectively. PROBAST + AI was used to systematically assess potential bias and concerns regarding applicability in prediction model studies through its structured checklist. QUADAS-2 is a widely used instrument for the quality appraisal of diagnostic test accuracy studies, providing an evaluation of both risk of bias and applicability across key methodological domains.

### 2.5 Data synthesis

LLMs performance was quantitatively synthesized through confusion matrix parameters (true positive (TP), false positive (FP), false negative (FN) and true negative (TN)). Where possible, unreported values were calculated using performance metrics (sensitivity and specificity).

To explore potential sources of substantial heterogeneity, subgroup analyses were conducted across six predefined dimensions: number of datasets, screening target, full-text screening, use of LLM-assisted techniques, model type, and prompt type. Meta-regression analyses were primarily performed based on four core dimensions, including number of datasets, screening target, full-text screening, and use of LLM-assisted techniques, which were available across all of included studies.

Due to data availability constraints, analyses regarding model type and prompt type were restricted to studies that explicitly reported these characteristics. Therefore, subgroup and meta-regression analyses for these two dimensions were conducted on a subset of eligible studies.

### 2.6 Statistical analysis

Diagnostic accuracy parameters, including sensitivity and specificity, were jointly estimated using a bivariate random-effects model and the hierarchical summary receiver operating characteristic (HSROC) framework. Data were summarized separately for studies evaluating full-text screening outcomes and those assessing title or abstract screening outcomes. Predefined subgroup analyses and meta-regression analyses were conducted using R software (version 4.2.1) and Stata (version 14.1). Statistical heterogeneity across studies was assessed using Cochran’s Q test and the I² statistic, which quantifies the proportion of total variability attributable to between-study heterogeneity rather than sampling error. An I² value below 50% was considered indicative of low heterogeneity, in which case a fixed-effects model was applied, whereas substantial heterogeneity (I² > 50%) warranted the use of a random-effects model to account for between-study variability.

## 3. Results

### 3.1 Study selection

A total of 3,192 records were identified across databases. After removal of 652 duplicate records, 2,540 unique records underwent title and abstract screening, of which 2,503 were excluded. Thirty-seven articles were retrieved for full-text assessment, and 19 were excluded due to the absence of diagnostic outcomes (n = 17) or lack of LLM-based methods (n = 2). Eighteen studies met the inclusion criteria and were included in both the systematic review and the meta-analysis (12, 17-33) (Fig. 1).

**Figure 1.**
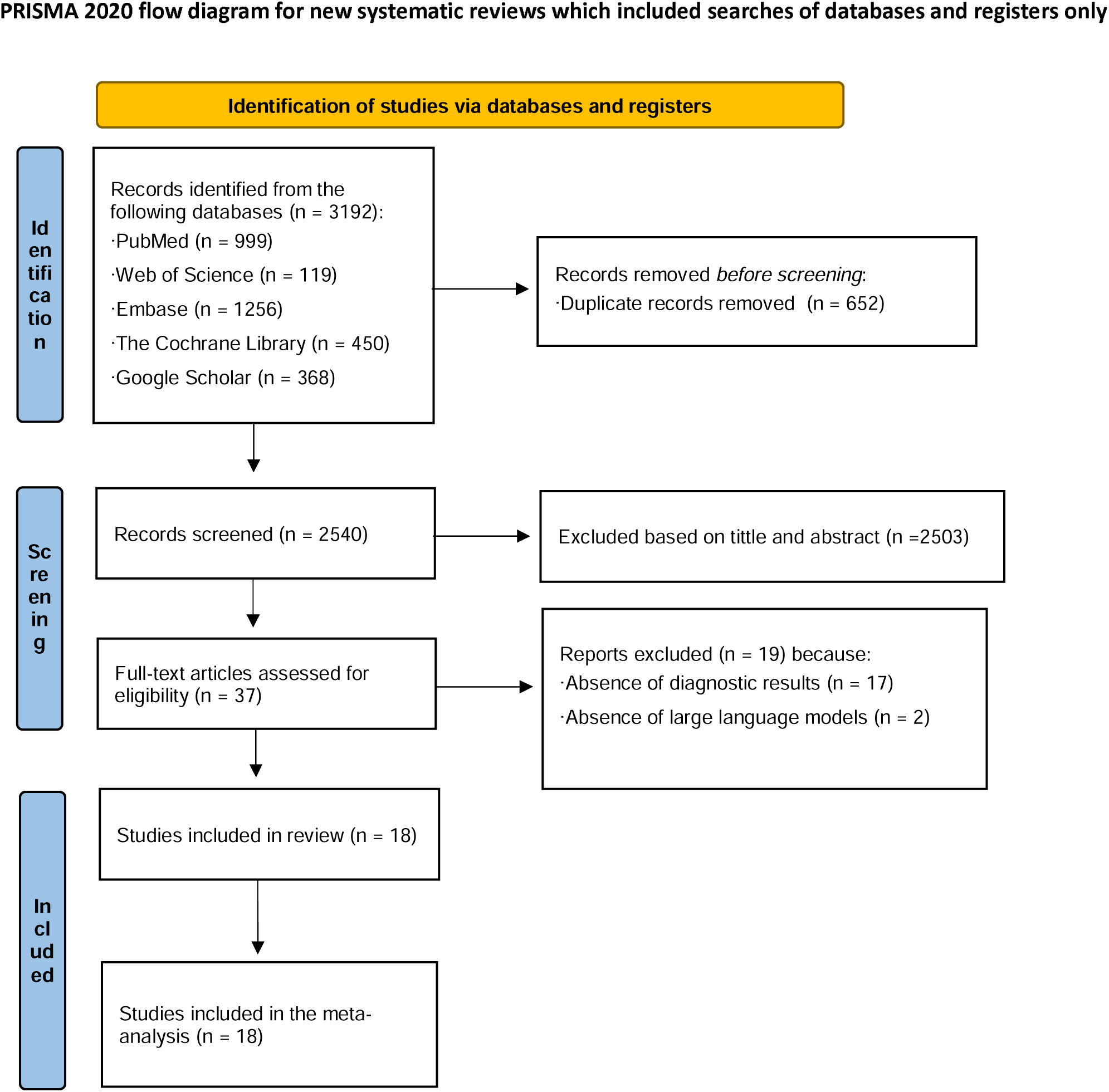
Preferred Reporting Items for Systematic reviews and Meta-Analyses (PRISMA) flowchart of study selection.

### 3.2 Study and model basic information

Table 1 summarizes the basic characteristics of the studies included in this systematic review on the performance of LLMs in automated medical literature screening. Overall, the included evidence comprised a diverse set of study designs, including systematic reviews, meta-analyses, scoping reviews, living systematic reviews, and original methodological or diagnostic accuracy studies. All studies were published between 2023 and 2025. Across studies, the number of embedded screening tasks varied substantially, ranging from single-review evaluations to large-scale experiments incorporating up to nine independent datasets within a single publication. The scope of screening predominantly focused on title and abstract screening, although several studies additionally evaluated full-text screening, either alone or in combination with earlier screening stages. Traditional manual screening by human reviewers was the most common gold standard, while a small number of studies incorporated hybrid or integrated artificial intelligence-assisted workflows as comparators. The volume of literature subjected to screening showed marked heterogeneity, from small datasets with fewer than 100 records to very large corpora exceeding 10,000 citations. Correspondingly, the number of truly included (eligible) studies after screening was generally low relative to the initial screening pool, often below 1-5% of the total records.

Table 2 presents the methodological characteristics and performance metrics of the LLM-based screening models evaluated in the included studies. A wide range of LLMs were assessed, including multiple generations of GPT-based models, Claude series models, Gemini models, LLaMA variants, and region-specific or domain-adapted LLMs. Most models were deployed via API-based access, while a smaller proportion relied on web-based interfaces. Importantly, the majority of studies used off-the-shelf LLMs without task-specific fine-tuning, emphasizing prompt engineering as the primary optimization strategy.

Prompting strategies varied considerably and included zero-shot prompting, few-shot prompting, chain-of-thought reasoning, role-promoting instructions, and criteria embedding. Several studies explicitly compared different prompting paradigms or ensemble strategies (e.g., majority voting or random forest-based aggregation) to improve screening performance. Model performance was typically evaluated against traditional manual screening or alternative automated tools, with some studies additionally incorporating internal methodological controls or comparisons with classical machine learning and natural language processing models.

In terms of diagnostic performance, reported sensitivity values for title and abstract screening were generally high but variable, ranging from moderate levels in conservative zero-shot settings to near-perfect sensitivity in optimized or ensemble-based configurations. Beyond diagnostic accuracy, many studies emphasized efficiency-related outcomes, including workload reduction, screening burden, time to decision, automation level, and cost savings. LLM-assisted screening consistently demonstrated substantial reductions in manual workload, frequently exceeding 60-90%, alongside dramatic decreases in screening time from hundreds of hours to a few hours or minutes.

### 3.3 Quality assessment

Table 3 summarizes the quality, applicability, and risk of bias of the included studies based on the PROBAST + AI tool assisted literature screening. Overall, the methodological quality was high across most studies, particularly in the development phase. The majority of studies demonstrated high quality in participant selection, predictor definition, outcome specification, and analytical methods during model development. Evaluation phases were also generally well conducted, although several studies showed reduced quality in analytical rigor. In terms of applicability, most studies exhibited low concern across both development and evaluation sections and a small number of studies showed reduced applicability. Regarding risk of bias, most studies were judged to have a low overall risk, particularly in the domains of predictors and outcomes. However, the analysis domain emerged as the most common source of high or unclear risk.

Table 4 showed the risk of bias and applicability by using the QUADAS 2 tool. Overall, most studies demonstrated a low risk of bias. The domains of reference standard and flow and timing were consistently rated as low risk across the majority of studies. In contrast, uncertainty was more frequently observed in the patient selection and index test domains. Applicability concerns were generally low. Most studies used screening tasks, datasets, and outcome definitions that closely resembled real world systematic or scoping review workflows. High applicability concern was identified in only one study, primarily related to patient selection, where the literature corpus was narrowly defined.

### 3.4 Meta-analysis

The meta-analysis evaluated the diagnostic performance of LLMs in automated medical literature screening by synthesizing data from the 18 studies. In automated title and abstract screening, the pooled sensitivity was 0.92 with 95% confidence interval (95% CI) from 0.81 to 0.96. Substantial heterogeneity was observed across studies with an I² value of 95.82. The pooled specificity was 0.94 with 95% CI from 0.90 to 0.97. And heterogeneity in specificity estimates was considerable with an I² value of 99.90 (Fig. 2). Moreover, the positive likelihood ratio (PLR) was 16.6 with 95% CI from 9.0 to 30.5 and the negative likelihood ratio (NLR) was 0.09 with 95% CI from 0.04 to 0.20. The diagnostic odds ratio (DOR) reached 185 with 95% CI from 68 to 503.

**Figure 2.**
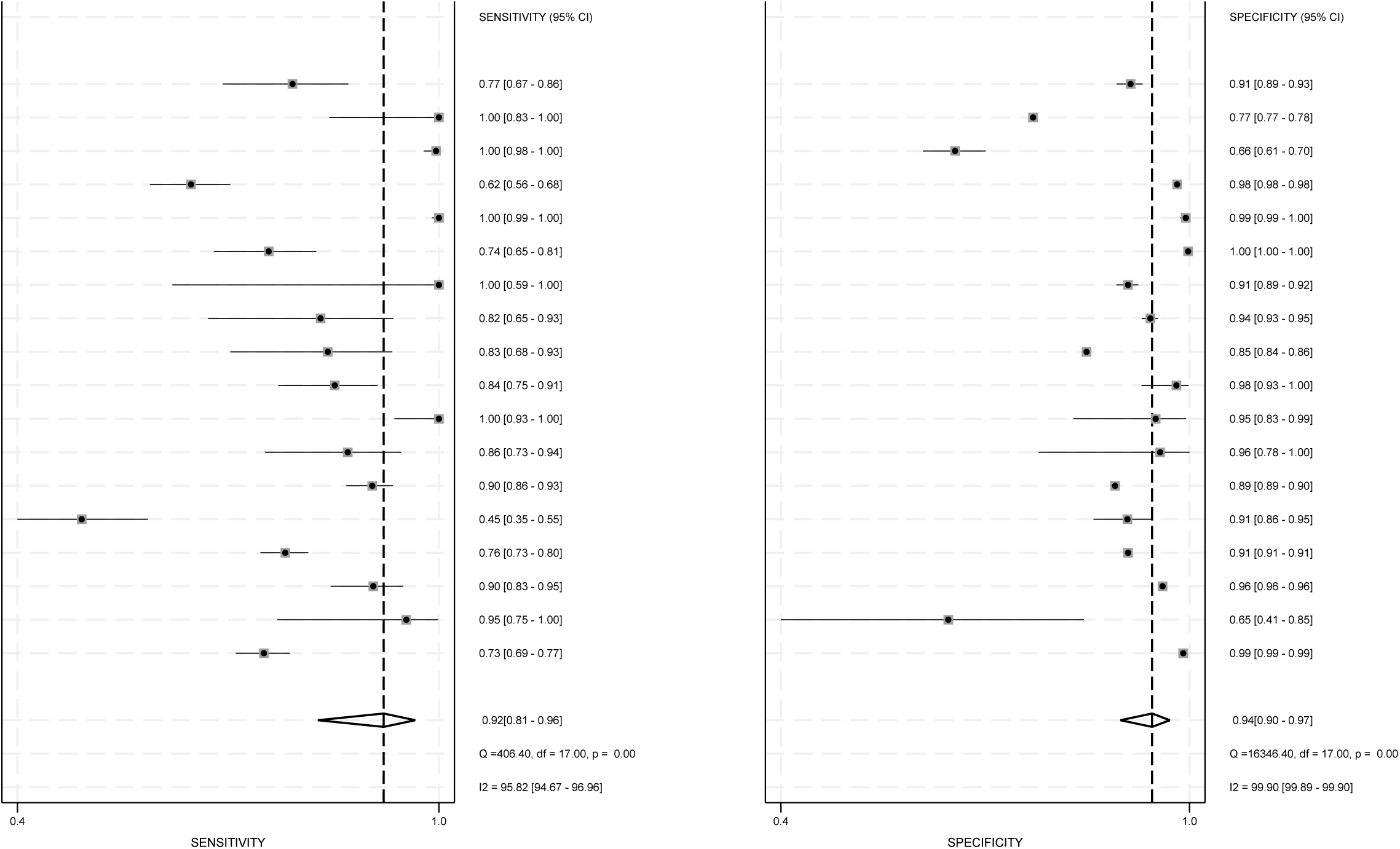
Forest plot of the random model meta-analysis of pooled sensitivity and specificity for automated title and abstract screening.

Summary receiver operating characteristic (SROC) analysis further confirmed the overall diagnostic performance. The area under the curve (AUC) reached 0.98 with 95% CI from 0.96 to 0.99 (Fig. 3).

**Figure 3.**
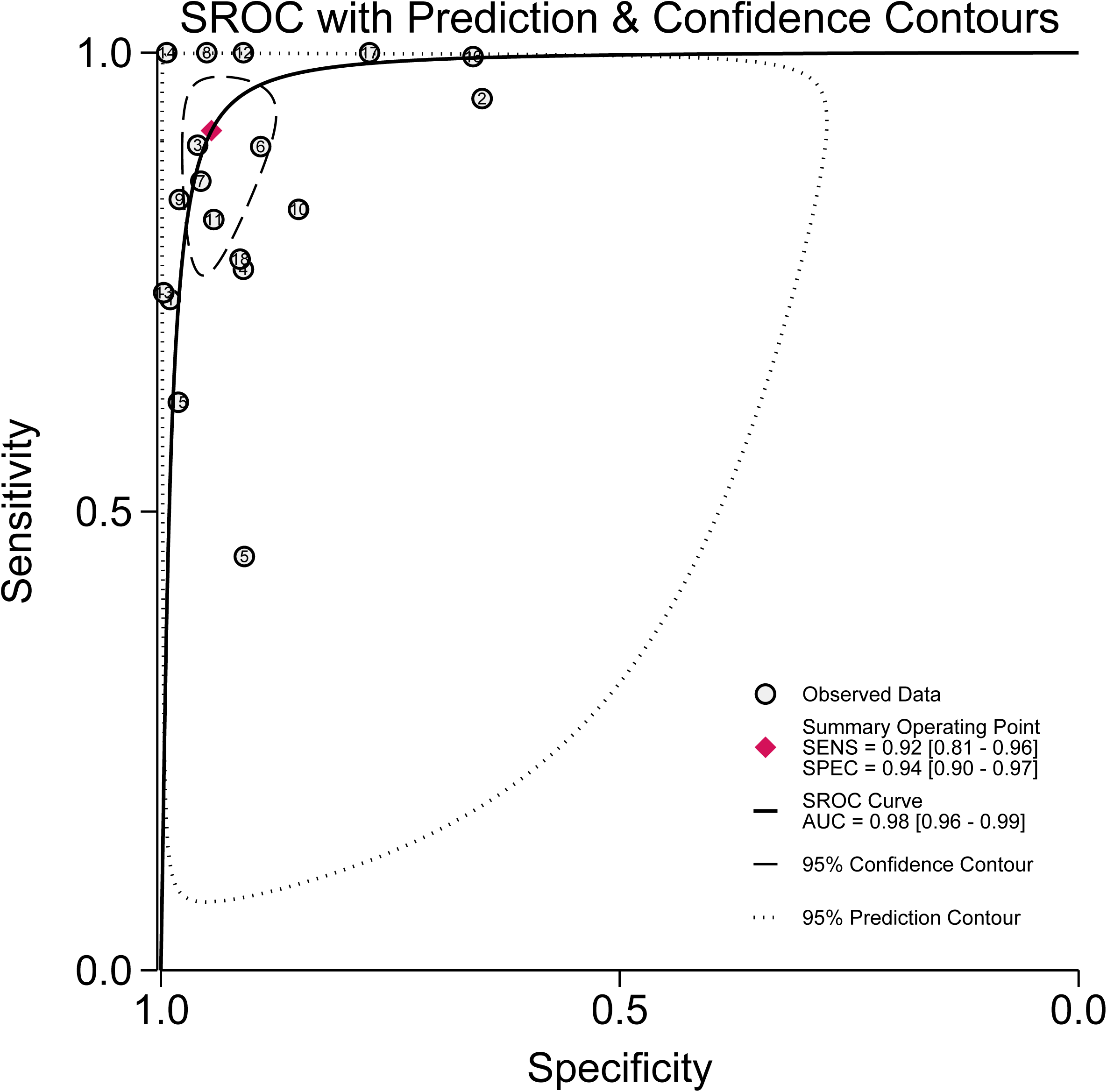
Receiver operating characteristics curves of the random model meta-analysis of pooled area under the curve for automated title and abstract screening.

Assessment of small study effects did not indicate evidence of publication bias. Deeks’ funnel plot asymmetry test showed no significant asymmetry with a bias coefficient of negative 1.16 and P value of 0.97 (Suppl. 2).

The meta-analysis of full-text screening included 4 studies. The pooled sensitivity reached 0.99 with 95% CI from 0.95 to 1.00. No substantial heterogeneity was detected with I² value of 0. The pooled specificity was 0.99 with 95% CI from 0.95 to 1.00. Considerable heterogeneity was observed in specificity estimates with I² value of 92.65 (Fig. 4). Moreover, the PLR was 77.2 with 95% CI from 17.6 to 338.5 and the NLR was 0.01 with 95% CI from 0 to 0.06. The DOR reached 9831 with 95% CI from 851 to 113516.

**Figure 4.**
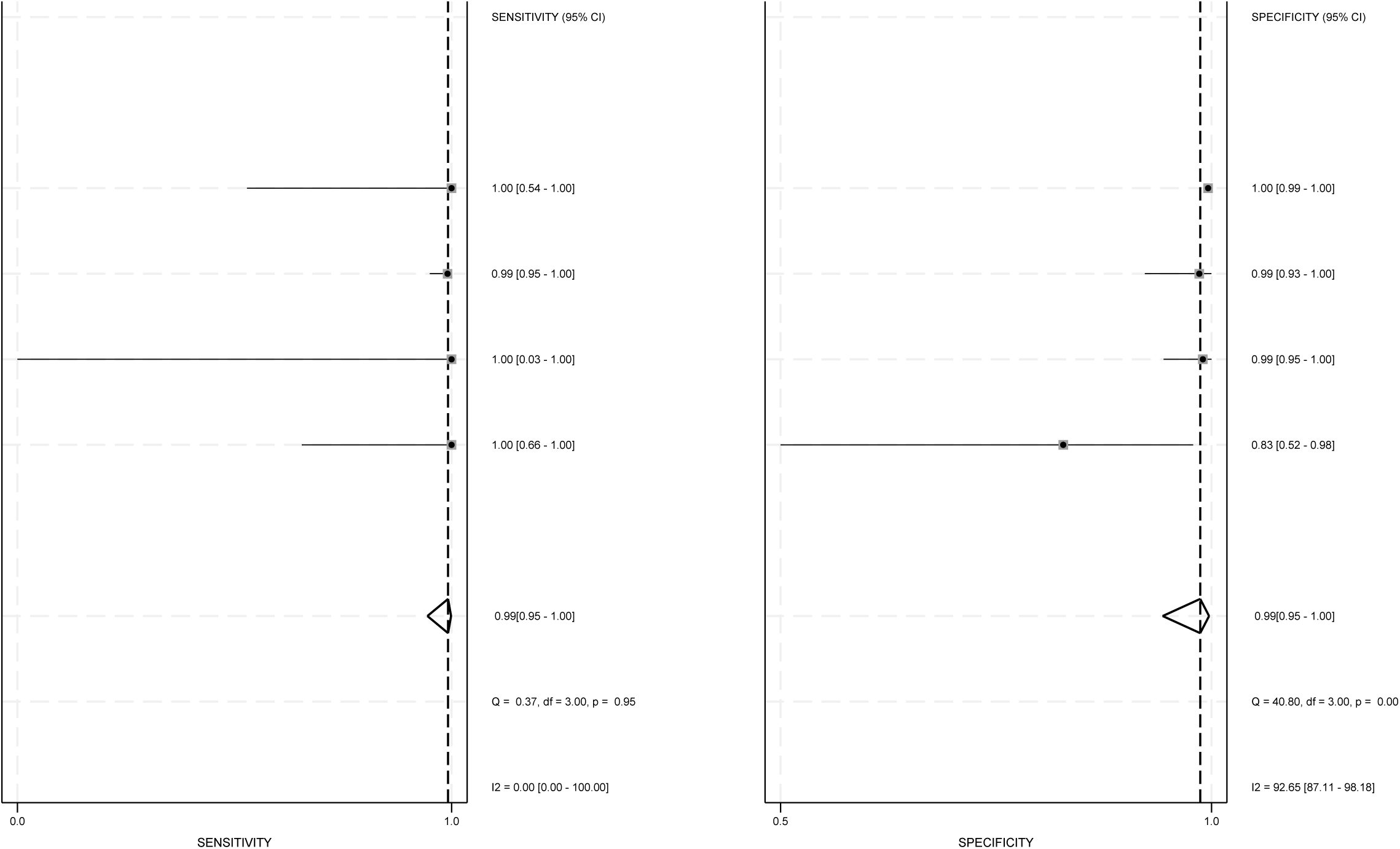
Forest plot of the random model meta-analysis of pooled sensitivity and specificity for automated full-text screening.

SROC analysis demonstrated strong diagnostic performance. The AUC reached 0.99 with 95% CI from 0.98 to 1.00 (Fig. 5).

**Figure 5.**
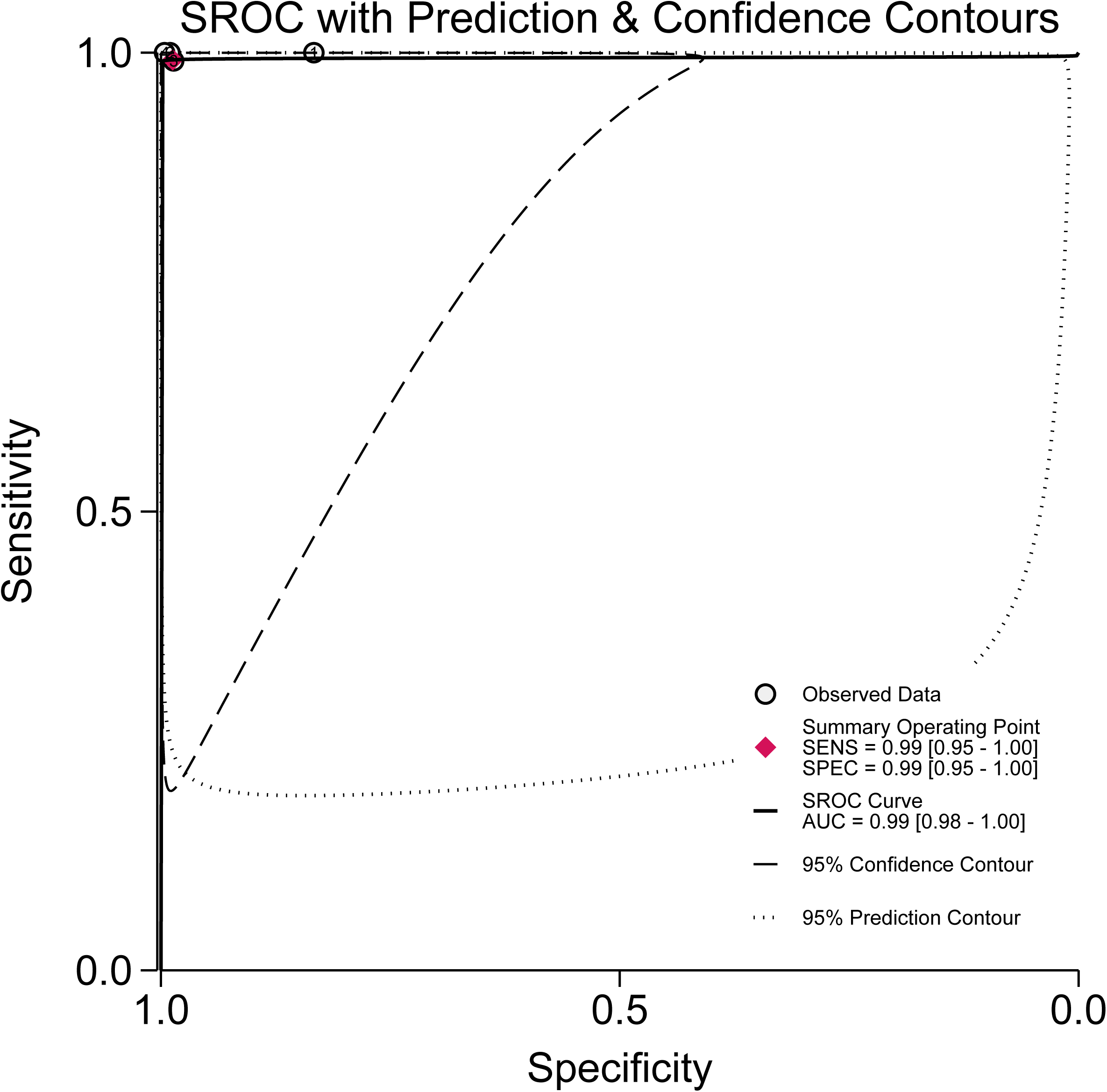
Summary receiver operating characteristic curves of the random model meta-analysis of pooled area under the curve for automated full-text screening.

Assessment of small study effects did not indicate clear evidence of publication bias. Deeks’ funnel plot asymmetry test showed no statistically significant asymmetry with a bias coefficient of negative 17.74 and P value of 0.17 (Suppl. 3).

### 3.5 Subgroup analysis and meta-regression

For full-text screening, studies that evaluated full-texts demonstrated a pooled sensitivity of 0.95 and specificity of 0.93, whereas studies focusing on non-full-text screening achieved a sensitivity of 0.89 and specificity of 0.95. The between subgroup differences were not statistically significant for either sensitivity or specificity, with p values of 0.36 and 0.10, respectively. In the joint model, the likelihood ratio test (LRT) did not indicate a significant covariate effect. With respect to dataset scale, studies using small datasets yielded a pooled sensitivity of 0.87 and specificity of 0.94, compared with 0.94 and 0.95 in studies using larger datasets. Although sensitivity appeared numerically lower in small dataset studies, the differences were not statistically significant, and meta regression confirmed the absence of a significant effect. Similarly, no meaningful differences were observed between systematic review and non-systematic review screening contexts. Sensitivity and specificity were 0.93 and 0.95 in systematic review settings, and 0.90 and 0.93 in other settings, with non-significant p values for both parameters. The joint model analysis further supported the lack of a significant subgroup effect. For studies incorporating assistive technology, pooled sensitivity and specificity were 0.94 and 0.96, compared with 0.90 and 0.94 in studies without such support. Despite slightly higher point estimates in the assistive technology subgroup, statistical testing did not demonstrate significant differences. LRT across all four covariates were non-significant, and residual heterogeneity within subgroups was minimal (Fig. 6).

**Figure 6.**
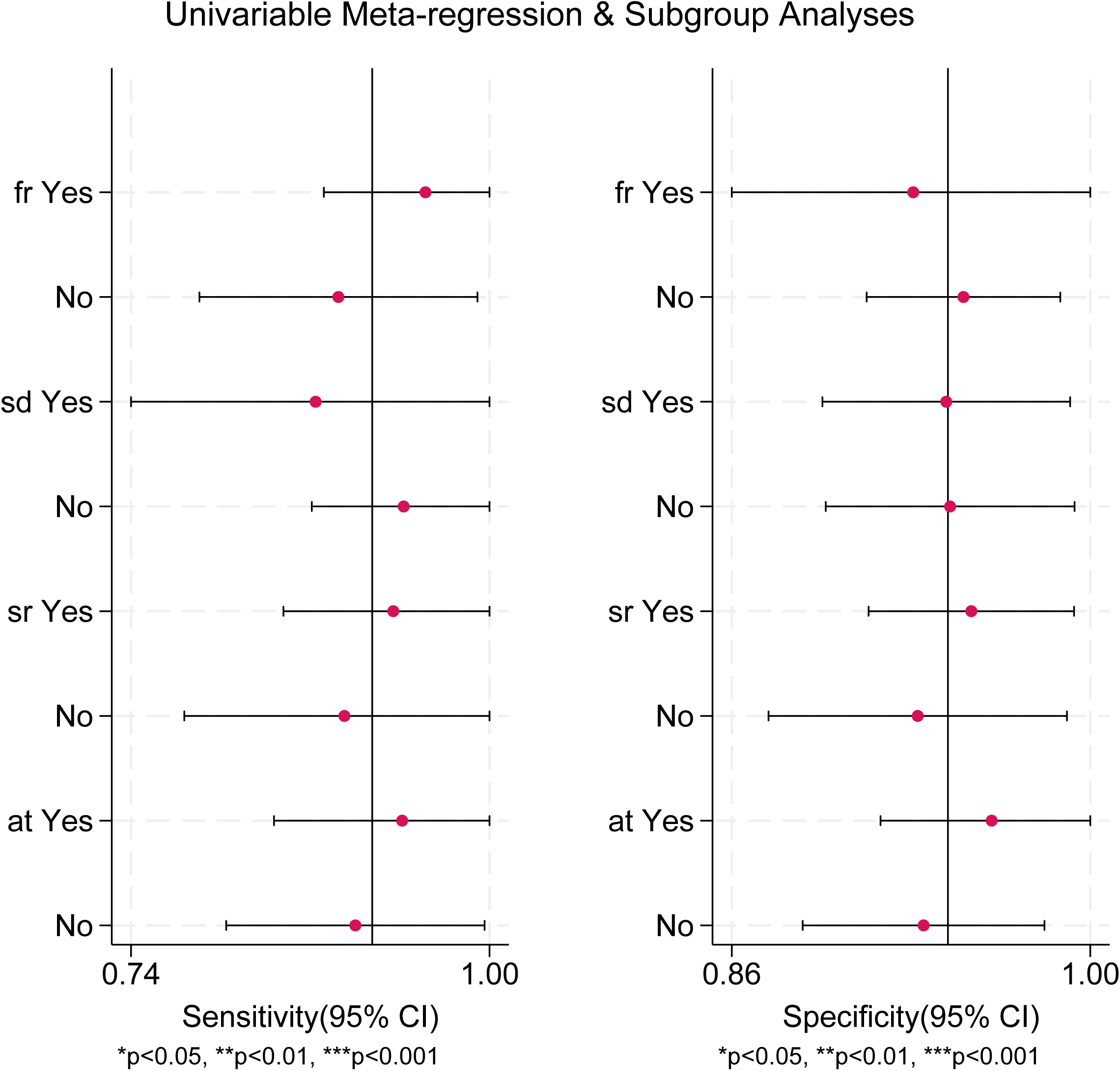

For model type, four studies evaluated GPT-4T and five evaluated GPT-4o. The pooled sensitivity was 0.81 with 95% CI from 0.69 to 0.93 for GPT-4T and 0.88 from 0.82 to 0.95 for GPT-4o. The difference in sensitivity was not statistically significant, with p value of 0.45. In contrast, specificity differed between groups. GPT-4T achieved a pooled specificity of 0.99 from 0.97 to 1.00, whereas GPT-4o reached 0.94 from 0.88 to 0.99, with p value < 0.01. However, the joint LRT for the covariate effect was not statistically significant, with p value of 0.13, and moderate residual heterogeneity was observed. For prompt strategy, four studies incorporated example-based or chain-of-thought prompting, whereas eight studies used neither example-based prompts nor chain-of-thought strategies. The pooled sensitivity was 0.95 with 95% CI from 0.92 to 0.99 in the example-based or chain-of-thought group and 0.86 from 0.78 to 0.94 in the group without these strategies. The difference in sensitivity was statistically significant, with p value < 0.01. Specificity was 0.91 from 0.79 to 1.00 in the example-based or chain-of-thought group and 0.98 from 0.96 to 1.00 in the group without these strategies, without a statistically significant difference. And the joint model yielded p value of 0.07, accompanied by substantial heterogeneity.

## 4 Discussion

In this systematic review and meta-analysis, we provide a comprehensive quantitative synthesis of the diagnostic performance of LLMs in automated medical literature screening. In 18 studies, LLMs shown high sensitivity and specificity in title and abstract screening, with AUC of 0.98. Performance was even stronger in full-text screening, where pooled sensitivity, specificity and AUC approached 0.99. The high sensitivity observed in title and abstract screening likely reflects the capability of LLMs to perform semantic reasoning. Unlike traditional rule-based approaches or conventional ML methods, LLMs can model semantic relationships through vector representations, enabling them to infer implicit inclusion and exclusion criteria as well as contextual associations embedded within titles and abstracts. At the same time, specificity remains high. Exclusion signals are often explicit in title and abstract sections, which makes the identification of clearly irrelevant records comparatively straightforward. In addition, when evidence is insufficient, LLMs tend to exclusion, a behavioral tendency that may further contribute to raise specificity. Similarly, full-text screening shown near perfect sensitivity and specificity. As full-text articles provide richer contextual information and more explicit inclusion and exclusion criteria information. LLMs appear particularly effective when supplied with sufficient semantic detail, enabling more reliable identification between eligible and ineligible studies(34).

The forest plots showed highly heterogeneity for both sensitivity and specificity across studies evaluating title and abstract screening. In contrast, in full-text screening studies, no heterogeneity was observed for sensitivity, whereas specificity contributed to considerable heterogeneity. We conducted predefined subgroup analyses and meta-regression to explore potential sources of this variability; however, no statistically significant contributors to heterogeneity were identified. On this basis, we observed that screening conditions across different settings may result in some influence on performance. Whether full-text screening was conducted had limited impact on overall results. Studies incorporating full-text screening showed slightly higher sensitivity, whereas non-full-text screening shown slightly higher specificity(35). With respect to dataset scale, larger datasets were associated with improved sensitivity, while specificity was comparable between larger and smaller datasets(36). Regarding screening targets, studies conducted within systematic review contexts had slightly higher sensitivity and specificity than those performed in non-systematic review settings(37). A similar fact was observed for assistive technology. Models implemented with assistive support achieved numerically higher sensitivity and specificity, suggesting a potential advantage of assisted workflows(38, 39). In comparisons between GPT-4o and GPT-4T, GPT-4o demonstrated higher sensitivity than GPT-4T, although the difference was not statistically significant. Conversely, GPT-4o showed lower specificity than GPT-4T, and this difference reached statistical significance. These findings indicate directional differences between model types, although the overall model level effect remains unstable(40, 41). With respect to prompt strategy, the example-based or chain-of-thought group achieved markedly higher sensitivity than the non-example and non-chain-of-thought group, and this difference was statistically significant(42). However, specificity was slightly lower in the example-based or chain-of-thought group, without a significant difference.

These findings have important application for medical evidence synthesis workflows. In title and abstract screening, the pooled sensitivity and specificity of LLMs both exceeded 0.90, while in full-text screening the pooled estimates approached unity. This level of performance suggests that LLMs are well suited for automated support across full stages of literature screening(43). In practical research settings, these AI tools can prioritize potentially relevant studies for human verification while substantially reducing the burden of manual screening(44). Several studies reported that LLMs assisted workflows reduced screening workload by at least fifty percent, with some reporting reductions exceeding ninety five percent(45). This allows researchers to devote more attention to higher level analytical tasks rather than routine screening procedures. Efficiency gains were also evident in terms of time and cost(46). Screening and decision-making time was commonly reduced by three to five fold, and in some cases improvements reached an order of magnitude. The deployment cost of LLMs assisted screening was also markedly lower, often representing less than one tenth of the cost of manual screening. As model architectures continue to evolve, improvements in performance are likely to be accompanied by further reductions in time and cost(47). LLMs are not intended to replace human reviewers. Instead, they function as assistive tools within the screening framework to streamline systematic review processes(48). Large scale evidence synthesis organizations are increasingly exploring automated approaches to optimize evidence production. Standardized integration of LLMs into literature screening and review management platforms may support rapid evidence updating while maintaining methodological rigorism(49).

Compared with earlier AI approaches for literature screening, which were primarily based on traditional text mining and classical ML algorithms, most supervised frameworks required labeled training datasets and extensive feature engineering(50, 51). These methods often depended on manually constructed features and validation procedures, which relied heavily on human effort. In specific medical domains, these models could achieve high sensitivity when trained on carefully annotated corpora. However, once applied to new medical topics or research areas, additional training data and redesigned feature pipelines were typically required(52). This substantially limited their generalizability and scalability across different evidence synthesis contexts. Another limitation of traditional approaches lies in their restricted ability for semantic understanding and reasoning(53). These systems often struggle to capture contextual relationships or infer implicit medical information. As a result, they are less capable of identifying nuanced eligibility criteria or complex methodological signals in titles and abstracts(54). In contrast, most studies included in this review evaluated off the shelf LLMs, with only a small number involving task specific fine-tuning. Despite minimal adaptation, overall diagnostic performance remained high. This finding suggests that general purpose LLMs possess transferable semantic reasoning capabilities that enable effective application to medical literature screening tasks.

This study has several limitations. First, substantial heterogeneity was observed across studies, particularly for specificity in title and abstract screening. Although subgroup analyses and meta-regression were conducted to explore potential sources of variability, the origins of heterogeneity were not fully explained and residual variation remained. Second, most included studies were methodological evaluations rather than real-world implementations. Third, the number of studies assessing full-text screening was limited, which may restrict the generalizability of the findings. Fourth, performance estimates relied on human defined reference standards, which are themselves subject to error and variability. Finally, incomplete reporting of confusion matrix data in some studies required reconstruction from summary metrics, which may introduce minor imprecision in the pooled estimates.

## 5 Conclusion

LLMs demonstrate high diagnostic accuracy in automated medical literature screening, with particularly strong performance in full-text assessment. Prompt strategies that incorporate examples or chain-of-thought reasoning can significantly improve screening sensitivity. Across models, however, different general purpose systems show inconsistent performance. LLMs are therefore best positioned as high sensitivity tools for medical literature screening that assist rather than replace human judgment. Continued methodological refinement and validation in large scale real-world settings will be essential to establish their long term role in evidence-based medicine.

## Data Availability

All data produced in the present work are contained in the manuscript

## Availability of Data and Materials

All data generated or analyzed during this study are included in this published article.

## Author Contributions

Chenggong Xie: conceptualization, investigation, data curation, methodology, formal analysis, and writing – original draft. Weichang Kong and Liuting Pi: investigation and data curation. Diaoxin Qi and Yuxuan Yang: formal analysis and validation. Hao Liang: methodology, supervision, and writing – review and editing. Bin Wang: supervision, project administration, and writing – review and editing. All authors approved the submission and take responsibility for the manuscript.

## Ethics Approval and Consent to Participate

Not applicable.

## Acknowledgment

Not applicable.

