## Supplementary Material 1 for "Performance of Large Language Models in Automated Medical Literature Screening: A Systematic Review and Meta-analysis"

Supplementary Material 1. Full details of the search strategies.

PubMed: ("ChatGPT"[Title/Abstract] OR "GPT"[Title/Abstract] OR "Generative pre-trained transformer"[Title/Abstract] OR "LLM"[Title/Abstract] OR "large language model"[Title/Abstract] OR "large language models"[Title/Abstract] OR "Generative language model"[Title/Abstract] OR "Gemini"[Title/Abstract] OR "Bard"[Title/Abstract] OR "Claude"[Title/Abstract] OR "Copilot"[Title/Abstract] OR "LLaMA"[Title/Abstract] OR "Deepseek"[Title/Abstract] OR "Qwen"[Title/Abstract] OR "AI"[Title/Abstract] OR "artificial intelligence"[Mesh] OR "artificial intelligence"[Title/Abstract] OR "machine learning"[Mesh] OR "machine learning"[Title/Abstract] OR "neural network"[Title/Abstract] OR "chatbot"[Title/Abstract] OR "transformer model"[Title/Abstract]) AND ("literature screening"[Title/Abstract] OR "study selection"[Title/Abstract] OR "systematic review screening"[Title/Abstract] OR "title screening"[Title/Abstract] OR "abstract screening"[Title/Abstract] OR "citation screening"[Title/Abstract] OR "evidence screening"[Title/Abstract] OR "screening automation"[Title/Abstract] OR "automated screening"[Title/Abstract] OR "machine learning screening"[Title/Abstract])

WOS: TI = ("ChatGPT" OR "GPT" OR "Generative pre-trained transformer" OR"LLM" OR "large language model" OR "large language models" OR"Generative language model" OR "Gemini" OR "Bard" OR"Claude" OR "Copilot" OR "LLaMA" OR "Deepseek" OR "Qwen" OR"AI" OR "artificial intelligence" OR "machine learning" OR"neural network" OR "chatbot" OR "transformer model") AND TI = ("literature screening" OR "study selection" OR"systematic review screening" OR "title screening" OR "abstract screening" OR"citation screening" OR "evidence screening" OR"screening automation" OR "automated screening" OR "machine learning screening")

Embase: ('ChatGPT':ti,ab OR 'GPT':ti,ab OR 'Generative pre-trained transformer':ti,ab OR 'LLM':ti,ab OR 'large language model':ti,ab OR 'large language models':ti,ab OR 'Generative language model':ti,ab OR 'Gemini':ti,ab OR 'Bard':ti,ab OR 'Claude':ti,ab OR 'Copilot':ti,ab OR 'LLaMA':ti,ab OR 'Deepseek':ti,ab OR 'Qwen':ti,ab OR 'AI':ti,ab OR 'artificial intelligence'/exp OR 'artificial intelligence':ti,ab OR 'machine learning'/exp OR 'machine learning':ti,ab OR 'neural network':ti,ab OR 'chatbot':ti,ab OR 'transformer model':ti,ab) AND ( 'literature screening':ti,ab OR 'study selection':ti,ab OR 'systematic review screening':ti,ab OR 'title screening':ti,ab OR 'abstract screening':ti,ab OR 'citation screening':ti,ab OR 'evidence screening':ti,ab OR 'screening automation':ti,ab OR 'automated screening':ti,ab OR 'machine learning screening':ti,ab)

Cochrane Library: (ChatGPT OR GPT OR "Generative pre-trained transformer" OR LLM OR "large language model" OR "large language models" OR "Generative language model" OR Gemini OR Bard OR Claude OR Copilot OR LLaMA OR Deepseek OR Qwen OR AI OR "artificial intelligence" OR "machine learning" OR "neural network" OR chatbot OR "transformer model") AND ("literature screening" OR "study selection" OR "systematic review screening" OR "title screening" OR "abstract screening" OR "citation screening" OR "evidence screening" OR "screening automation" OR "automated screening" OR "machine learning screening" OR screening)

Google Scholar: ("LLM" OR "large language model") AND ("systematic review" OR "meta-analysis") AND ("automated screening" OR "screening automation" OR "machine learning screening")
