## Supplementary figures and images for "Performance of Large Language Models in Automated Medical Literature Screening: A Systematic Review and Meta-analysis"

### Supplementary Material 2

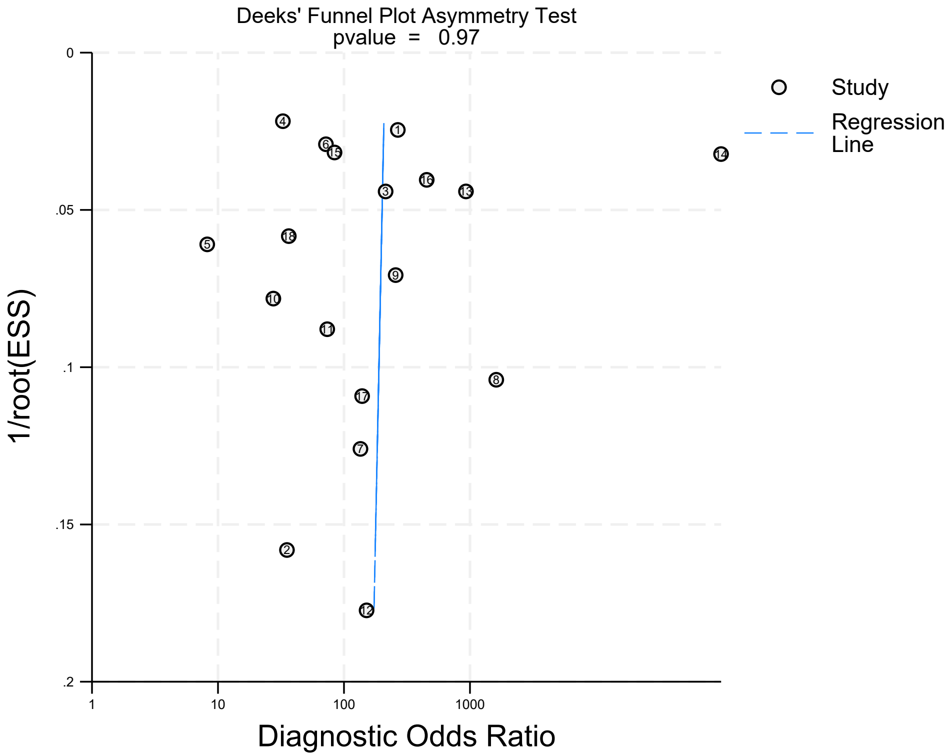

### Supplementary Material 3

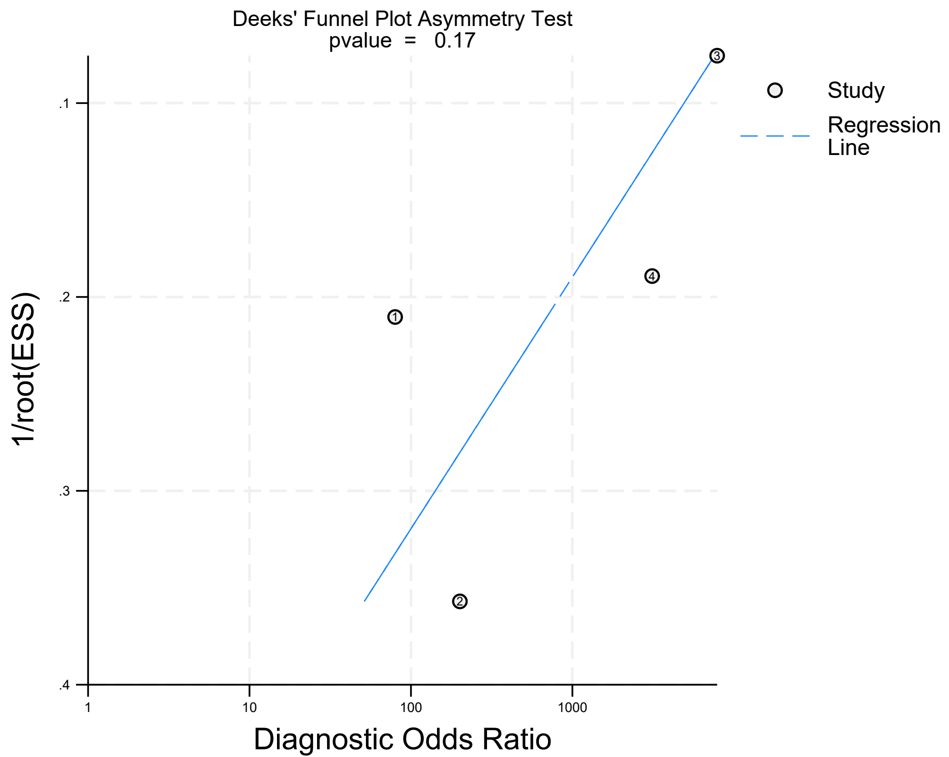
