## Supplementary material for "Performance of Large Language Models in Automated Medical Literature Screening: A Systematic Review and Meta-analysis": Table 1

Table 1. The basic characteristics of the studies included in this systematic review.

| **Title** | **First author**  **(year)** | **Types of studies included** | **Number of studies included** | **Retrieve content** | **Gold standard establishment method** | **The number of literature to be screened** | **The number of actually included literature** |
| --- | --- | --- | --- | --- | --- | --- | --- |
| Is Large Language Model-Assisted Citation Screening Feasible in a Scoping Review on Nonpharmacological Interventions for Delirium in Patients With Cancer? | Yoshiyasu Ito (2025) | Meta-analysis | Not mentioned | Title and abstract screening, full text screening | Traditional manual screening | Not mentioned | Not mentioned |
| Large language models for abstract screening in systematic- and scoping reviews: A diagnostic test accuracy study | Christian Hedeager Krag (2024) | Systematic reviews, scoping review | 2 | Abstract screening | Traditional manual screening | study1: 500 study2: 500 | study1: 12 study2: 130 |
| Evaluating the efectiveness of large language models in abstract screening: a comparative analysis | Michael Li (2024) | Systematic reviews | 3 | Abstract screening | Traditional manual screening | study1: 1488 study2: 969 study3: 631 | Not mentioned |
| Development and evaluation of prompts for a large language model to screen titles and abstracts in a living systematic review | Ava Homiar (2025) | Living systematic reviews | 3 | Title and abstract screening, full text screening | Traditional manual screening | study1: 11939 study2: 167 study3: 334 | study1: 60 study2: 2 study3: 2 |
| Sensitivity, specificity and avoidable workload of using a large language models for title and abstract screening in systematic reviews and meta-analyses | Viet-Thi Tran (2023) | Systematic reviews, meta-analysis | 5 | Title and abstract screening | Traditional manual screening | study1: 673 study2: 4077 study3: 6334 study4: 6478 study5: 5104 | study1: 26 study2: 8 study3: 200 study4: 19 study5: 179 |
| Accuracy of Large Language Models for Literature Screening in Thoracic Surgery: Diagnostic Study | Zhang-Yi Dai (2025) | Meta-analysis | 6 | Title and abstract screening | Traditional manual screening | study1: 357 study2: 462 study3: 296 study4: 429 study5: 2298 study6: 696 | study1: 28 study2: 12 study3: 26 study4: 11 study5: 10 study6: 26 |
| Automated Literature Screening for Hepatocellular Carcinoma Treatment Through Integration of 3 Large Language Models: Methodological Study | Chen Pan (2025) | Meta-analysis | 9 | Title and abstract screening | Integrating artificial intelligence | study1: 770 study2: 5417 study3: 309 study4: 1698 study5: 8733 study6: 558 study7: 897 study8: 3522 study9: 6078 | study1: 11 study2: 11 study3: 12 study4: 9 study5: 15 study6: 21 study7: 13 study8: 13 study9: 24 |
| LitAutoScreener: Development and Validation of an Automated Literature Screening Tool in Evidence-Based Medicine Driven by Large Language Models | Yiming Tao (2025) | Systematic reviews | 4 | Title and abstract screening, full-text screening | Traditional manual screening | Not mentioned | Not mentioned |
| Automated Paper Screening for Clinical Reviews Using Large Language Models: Data Analysis Study | Eddie Guo (2024) | Original research | 6 | Title and abstract screening |  | study1: 279 study2: 3989 study3: 1456 study4: 942 study5: 14771 study6: 2870 | study1: 35 study2: 29 study3: 91 study4: 6 study5: 354 study6: 23 |
| Assessing the performance of large language models in literature screening for pharmacovigilance: a comparative study | Dan Li (2024) | Original research | 3 | Abstract screening | Traditional manual screening | study1: 30 study2: 40 study3: 200 | study1: 15 study2: 20 study3: 40 |
| Enhancing AI for citation screening in literature reviews: Improving accuracy with ensemble models | Zhihong Zhang (2025) | Original research | 3 | Title and abstract screening | Traditional manual screening | study1: 865 study2: 959 study3: 73 | study1: 431 study2: 61 study3: 50 |
| Evaluating GPT Models for Automated Literature Screening in Wastewater-Based Epidemiology | Kaseba Chibwe (2025) | Meta-analysis | 2 | Abstract screening | Traditional manual screening | Not mentioned | Not mentioned |
| GPT-3.5 Turbo and GPT-4 Turbo in Title and Abstract Screening for Systematic Reviews | Takehiko Oami (2025) | Systematic reviews | 1 | Title and abstract screening | Traditional manual screening | study1: 16669 | study1: 41 |
| Validation of large language models (Llama 3 and ChatGPT-4o mini) for title and abstract screening in biomedical systematic reviews | Adriana López-Pineda (2025) | Systematic reviews | 1 | Title and abstract screening | Traditional manual screening | study1: 1081 | study1: 84 |
| Performance of a Large Language Model in Screening Citations | Takehiko Oami (2024) | Systematic reviews | 5 | Title and abstract screening | Traditional manual screening | study1: 5634 study2: 3418 study3: 1038 study4: 4326 study5: 2253 | study1: 8 study2: 4 study3: 4 study4: 17 study5: 8 |
| Can large language models replace humans in systematic reviews? Evaluating GPT-4's efficacy in screening and extracting data from peer-reviewed and grey literature in multiple languages | Qusai Khraisha (2024) | Original research | 3 | Title and abstract screening, full text screening | Traditional manual screening | study1: 300 study2: 150 study3: 30 | Not mentioned |
| How Well Do ChatGPT and Claude Perform in Study Selection for Systematic Review in Obstetrics | Suppachai Insuk (2025) | Systematic reviews | 2 | Title and abstract screening, full text screening | Traditional manual screening | study1: 1648 study2: 21 | study2: 9 |
| Streamlining systematic reviews with large language models using prompt engineering and retrieval augmented generation | Fouad Trad (2025) | Systematic reviews | 2 | Title and abstract screening, full text screening | Traditional manual screening | study1: 14439 study2: 3298 | study2: 78 |
