## Supplementary material for "Performance of Large Language Models in Automated Medical Literature Screening: A Systematic Review and Meta-analysis": Table 2

Table 2. The methodological characteristics and performance metrics of the LLM-based screening models evaluated in the included studies.

| **Title** | **First author**  **(year)** | **The LLMs and its version** | **The LLMs deployment methods** | **Prompt type** | **Is the LLMs fine-tuned?** | **LLMs comparison method** | **LLMs performance metrics** | **LLMs efficiency metrics** |
| --- | --- | --- | --- | --- | --- | --- | --- | --- |
| Accuracy of Large Language Models for Literature Screening in Thoracic Surgery: Diagnostic Study | Zhang-Yi Dai (2025) | GPT-4o, Claude-3.5 and Gemini-1.5 pro | Web-based | Chain-of-thought, criteria Emending | No | Model intergroup comparison | 1. Title/Abstract: (1)Sensitivity: Pooled: 0.73 (95%CI 0.57–0.85) (2)Specificity: Pooled: 0.99 (95%CI 0.97–0.99) (3)AUC: 0.97 (95%CI 0.96–0.99) 2. Full Text: (1)Sensitivity: Study 1: 0.93 (95%CI 0.76–0.99); Study 2: 0.75 (95%CI 0.43–0.95); Study 3: 0.92 (95%CI 0.75–0.99); Study 4: 1.00 (95%CI 0.72–1.00); Study 5: 0.90 (95%CI 0.55–1.00); Study 6: 0.69 (95%CI 0.48–0.86); Pooled: 0.87 (95%CI 0.77–0.99) (2)Specificity: Study 1: 0.84 (95%CI 0.80–0.88); Study 2: 0.94 (95%CI 0.91–0.96); Study 3: 0.97 (95%CI 0.94–0.99); Study 4: 0.92 (95%CI 0.89–0.94); Study 5: 0.99 (95%CI 0.98–0.99); Study 6: 0.99 (95%CI 0.98–1.00); Pooled: 0.96 (95%CI 0.91–0.98) (3)AUC: 0.96 (95%CI 0.94–0.97) 3. Post-Hoc Analysis (Full Text): (1)Sensitivity: Pooled: 0.98 (95%CI 0.74–1.00) (2)Specificity: Pooled: 0.98 (95%CI 0.94–0.99) (3)AUC: 1.00 (95%CI 0.99–1.00) | 1.Sensitivity: Traditional machine learning-assisted screening achieved 0.24–0.80, while semi-automated tools achieved 0.35–0.64. 2.Time to decision: Reduced by tenfold. |
| Assessing the performance of large language models in literature screening for pharmacovigilance a comparative study | Dan Li (2024) | GPT-3.5, GPT-4 and Claude-2 | API | Chain-of-thought, zero shot, few-shot | No | Model intergroup comparison, developed BERT series models (NLP-DILI model) as control | 1.Sensitivity: Best 0.975, Median 0.9 2.Specificity: Median 0.67 3.Precision: Median 0.735 4.Accuracy: 0.7–0.8 5.F1-score: 0.694–0.83 | 1.Workload saved:70% 2.Screening burden: With theassist of LLMs,5,950 irrelevant abstracts (70% of 8,500)could be filtered out. |
| Automated Literature Screening for Hepatocellular Carcinoma Treatment Through Integration of 3 Large Language Models：Methodological Study | Chen Pan(2025) | Doubao-1.5-pro-32k, Deepseek-3 and DeepSeek-R1-Distill-Qwen-7 | API | Chain-of-thought, role-promoting, criteria embeding | No | Comparison with Research Tools(Scispace, Elicit, Consensus), Screening Tools(DistillerSR, RobotAnalyst) and GPT-4o | 1.Sensitivity (1)For Inclusion tasks: Weighted average 0.90 (SD 0.16; range 0.62–1.00) (2)For Exclusion tasks: Weighted average 0.96 (SD 0.04; range 0.90–1.00) 2.Precision (1)For Inclusion tasks: Weighted average 0.15 (SD 0.13; range 0.05–0.47) (2)For Exclusion tasks: Weighted average 1.00 (SD < 0.01; range 0.99–1.00) 3.Accuracy (1)Overall: 95.54% (2)Weighted average: 0.96 4.F1-score For Inclusion tasks: Weighted average 0.22 (SD 0.16; range 0.04–0.58) 5.κ (Kappa): Mean κ: 0.258 6.PABAK (1)Mean PABAK: 0.873 (2)Weighted mean PABAK: 0.91 (range 0.76–0.99) | 1. Workload Reduction Overall: In review updates, manual screening workload is reduced by nearly 80% without missing eligible studies. Specific Items: Baseline Review: Workload reduced by 65.72% Update 1: Workload reduced by 79.04% Update 2: Workload reduced by 85.63% 2. Time Savings: Approximately 260–650 hours of manual screening time saved. |
| Automated Paper Screening for Clinical Reviews Using Large Language Models：Data Analysis Study | Eddie Guo(2024) | GPT-4 | API | Zero-shot, role-promoting | No | Comparison to Other Tool(such as Abstrackr , DistillerSR, and RobotAnalyst ) | 1.Sensitivity: (1)For inclusion tasks0.764(95%CI 0.593 – 1.000)  (2)For exclusion tasks:0.910(95%CI 0.756 – 0.966) 2.Specificity:0.91  3.Accuracy:0.907(95%CI 0.748 – 0.965) 4.F1-score:0.6(95%CI 0.594 – 0.613) | 1.The model ran for 643 minutes and 50.8 seconds, with an approximate cost of US $25. 2.On the nonopioid analgesia (NOA) data set (354/14,771 included abstracts), the model ran for 643 minutes and 50.8 seconds. |
| Can large language models replace humans in systematic reviews？Evaluating GPT-4's efficacy in screening and extracting data from peer-reviewed and grey literature in multiple languages | QusaiKhraisha(2023) | GPT-4 | ChatGPT interface | Zero-shot, Criteria embedding | No | Internal methodological control | 1. Title/Abstract: (1)Sensitivity: ① English Peer-Reviewed: 0.42; ② English Grey Literature: 0.4; ③ Other Languages: 0.50 (2)Specificity: ① English Peer-Reviewed: 0.92; ② English Grey Literature: 0.84; ③ Other Languages: 0.89 (3)Accuracy: ① English Peer-Reviewed: 0.67; ② English Grey Literature: 0.66; ③ Other Languages: 0.88 2. Full-Text: (1)Sensitivity: ① English Peer-Reviewed: 0.38; ② English Grey Literature: 0.60; ③ Other Languages: 1.00; ④ High-Reliability Prompt Group: 0.36 (2)Specificity: ① English Peer-Reviewed: 0.69; ② English Grey Literature: 0.80; ③ Other Languages: 0.95; ④ High-Reliability Prompt Group: 0.94 (3)Accuracy: ① English Peer-Reviewed: 0.54; ② English Grey Literature: 0.78; ③ Other Languages: 0.96; ④ High-Reliability Prompt Group: 0.85 | not mentioned |
| Development and evaluation of prompts for a large language model to screen titles and abstracts in a living systematic review | Ava Homiar(2025) | GPT-4o | API | Zero-shot | No | Internal methodological control | 1.Title/Abstract:  (1)Sensitivity: ① Baseline: 0.91; ② Update 1: 1.00; ③ Update 2: 0.58 (2) Specificity: ① Baseline: 0.89; ② Update 1: 0.87; ③ Update 2: 0.92 (3) Precision: ① Baseline: no; ② Update 1: 0.04; ③ Update 2: 0.20 2.Full-text:  (1)Sensitivity: ① Baseline: 1.00; ② Update 1: 1.00; ③ Update 2: 1.00 (2) Specificity: ① Baseline: 0.88; ② Update 1: 0.87; ③ Update 2: 0.91 (3) Precision: ① Baseline: no; ② Update 1: 0.04; ③ Update 2: 0.04 | 1.Workload saved:(1)Baseline review: 65.72% workload reduction(2)Update 1: 79.04% workload reduction (3)Update 2: 85.63% workload reduction;(4)Overall conclusion: In review updates, LLMs can reduce manual screening workload by nearly 80% without missing eligible studies. 2.Screening burden:Baseline: 3,004 (vs. 11,939); Update 1: 35 (vs. 167); Update 2: 48 (vs. 334) 3.Time to decision:Manual screening: 2–5 min/record; total time reduced from 400–1,000 hrs to 140–350 hrs |
| Enhancing AI for citation screening in literature reviews：Improving accuracy with ensemble models | Zhihong Zhang(2025) | GPT-4o Mini, GPT-4o, Gemini Flash, Llama-3.1 8B Instruct, Llama-3.1 70B Instruct and Llama-3.1 405B Instruct | Not mentioned | Zero-shot, few-shot | No | Model intergroup comparison, comparison under few-shot vs. zero-shot scenarios, comparisons with three different review databases (LLM-Healthcare, MCI-Speech, Multimodal-LLM), and hierarchical comparison of predictive methods (single LLM prediction, majority vote ensemble, random forest ensemble). | 1.LLM-Healthcare Dataset (1) Best Model: GPT-4o (Random Forest Ensemble) ① Sensitivity: 0.96 (95%CI 0.93–0.99) ② Specificity: 0.89 (95%CI 0.84–0.93) (2) Average Performance of Individual LLMs ① Sensitivity Range: 0.84–0.98 ② Specificity Range: 0.55–0.83 2.MCI-Speech Dataset (1) Best Model (Majority Voting): Majority_vote_LLMs_1shot_1vote ① Sensitivity: 0.75 (95%CI 0.54–0.96) ② Specificity: 0.86 (95%CI 0.82–0.90) (2) Best Model (Random Forest Ensemble): LLMs-4shot ① Sensitivity: 0.62 (95%CI 0.39–0.86) ② Specificity: 0.97 (95%CI 0.96–0.99) (3) Average Performance of Individual LLMs ① Sensitivity Range: 0.44–0.62 ② Specificity Range: 0.84–0.97 3.Multimodal-LLM Dataset (1) Best Model: Multiple Random Forest Ensemble Models ① Sensitivity: 1.0 (95%CI 1.0–1.0) ② Specificity: 1.0 (95%CI 1.0–1.0) (2) Average Performance of Individual LLMs ① Sensitivity Range: 0.60–1.00 ② Specificity Range: 0.71–1.00 | Time to decision:an individual LLM processed one citation in just 0.5–1.5 s—far quicker than the time typically required by human reviewers. |
| EvaluatingGPTModelsforAutomatedLiteratureScreeningin Wastewater-BasedEpidemiology | KasebaChibwe(2024) | GPT-3, GPT-3.5T and GPT-4 | API | Zero-shot, few-shot | Yes  (GPT-3) | Model intergroup comparison, comparison before and after fine-tuning, and internal variable control | 1. Abstract Screening for Original Data Detection  (1) Sensitivity  ① GPT-4(Chat Completion): 1.00 ② GPT-3.5T(Chat Completion): 1.00 ③ text-davinci-003 [GPT-3] (Text Completion): 0.94  (2) Precision ① GPT-4(Chat Completion): 0.96 ② GPT-3.5T(Chat Completion): 0.72 (3) F1-Score ① GPT-4 (Chat Completion): 0.98 ② GPT-3.5T(Chat Completion): 0.89 ③ text-davinci-003 [GPT-3] (Text Completion): 0.94 2. Abstract Screening for Relevant Sampling Location Detection  (1) Sensitivity  ① Fine-tuned davinci [GPT-3] classifier: 0.84(SD = 0.04) ② Fine-tuned ada [GPT-3] classifier: 0.83(SD = 0.05) ③ GPT-4(Chat Completion): 0.60 ④ GPT-3.5T(Chat Completion): 0.43 (2) F1-Score ① Fine-tuned davinci [GPT-3] classifier: 0.83(SD = 0.04) ② Fine-tuned ada [GPT-3] classifier: 0.77(SD = 0.10) ③ GPT-4(Chat Completion): 0.66 ④ GPT-3.5T(Chat Completion): 0.60 | 1.Workload Reduction: reduce approximately 70% 2.Time Efficiency: a reduction of >95%  3.Low Cost: The cost per 100 abstracts screened is approximately $1 for GPT-4 and $0.07 for GPT-3.5-Turbo. |
| Evaluating the efectiveness of large language models in abstract screening: a comparative analysis | Michael Li(2024) | GPT-4, GPT-3.5, Google PaLM-2, Meta Llama-2, GPT-4T, GPT-3.5T, Gemini-1 pro, Meta Llama-3 and Claude-3 | API | Zero-shot, Role prompting | No | Model intergroup comparison, internal variable control. | 1.Title/Abstract（Bannach-Brown 2016） (1) Sensitivity ① ChatGPT (v4.0)：0.930 (95%CI 0.861, 0.971) ② ChatGPT (v3.5)：0.940 (95%CI 0.874, 0.978) ③ Google PaLM 2：0.850 (95%CI 0.765, 0.914) ④ Meta Llama 2：0.950 (95%CI 0.887, 0.984) ⑤ ChatGPT-3.5-Turbo：0.830 (95%CI 0.742, 0.898) ⑥ ChatGPT-4-Turbo：0.670 (95% 0.569, 0.761) ⑦ Gemini-1.0-pro：0.750 (95% 0.653, 0.831) ⑧ Llama 3：0.930 (95% 0.861, 0.971) ⑨ Claude 3 Opus：0.900 (95% 0.824, 0.951) (2) Specificity（95%CI） ① ChatGPT (v4.0)：0.960 (95% 0.901, 0.989) ② ChatGPT (v3.5)：0.870 (95% 0.788, 0.929) ③ Google PaLM 2：0.950 (95% 0.887, 0.984) ④ Meta Llama 2：0.610 (95% 0.507, 0.706) ⑤ ChatGPT-3.5-Turbo：0.910 (95% 0.836, 0.958) ⑥ ChatGPT-4-Turbo：0.990 (95% 0.946, 1.000) ⑦ Gemini-1.0-pro：0.990 (95% 0.946, 1.000) ⑧ Llama 3：0.890 (95% 0.812, 0.944) ⑨ Claude 3 Opus：0.940 (95% 0.874, 0.978) (3) Accuracy（95%CI） ① ChatGPT (v4.0)：0.945 (95% 0.904, 0.972) ② ChatGPT (v3.5)：0.905 (95% 0.856, 0.942) ③ Google PaLM 2：0.900 (95% 0.850, 0.938) ④ Meta Llama 2：0.780 (95% 0.716, 0.835) ⑤ ChatGPT-3.5-Turbo：0.870 (95% 0.815, 0.913) ⑥ ChatGPT-4-Turbo：0.830 (95% 0.771, 0.879) ⑦ Gemini-1.0-pro：0.870 (95% 0.815, 0.913) ⑧ Llama 3：0.910 (95% 0.861, 0.946) ⑨ Claude 3 Opus：0.920 (95% 0.873, 0.954) 2.Title/Abstract（Meijboom 2021） (1) Sensitivity（95%CI） ① ChatGPT (v4.0)：0.812 (95% 0.636, 0.928) ② ChatGPT (v3.5)：0.969 (95% 0.838, 0.999) ③ Google PaLM 2：0.647 (95% 0.383, 0.858) ④ Meta Llama 2：1.000 (95% 0.891, 1.000) ⑤ ChatGPT-3.5-Turbo：0.970 (95% 0.840, 0.999) ⑥ ChatGPT-4-Turbo：0.560 (95% 0.374, 0.734) ⑦ Gemini-1.0-pro：0.380 (95% 0.215, 0.568) ⑧ Llama 3：0.970 (95% 0.840, 0.999) ⑨ Claude 3 Opus：0.910 (95% 0.755, 0.982) (2) Specificity（95%CI） ① ChatGPT (v4.0)：0.860 (95% 0.776, 0.921) ② ChatGPT (v3.5)：0.470 (95% 0.369, 0.572) ③ Google PaLM 2：0.954 (95% 0.871, 0.990) ④ Meta Llama 2：0.520 (95% 0.418, 0.621) ⑤ ChatGPT-3.5-Turbo：0.570 (95% 0.467, 0.669) ⑥ ChatGPT-4-Turbo：0.930 (95% 0.861, 0.971) ⑦ Gemini-1.0-pro：0.960 (95% 0.901, 0.989) ⑧ Llama 3：0.870 (95% 0.788, 0.929) ⑨ Claude 3 Opus：0.840 (95% 0.753, 0.906) (3) Accuracy（95%CI） ① ChatGPT (v4.0)：0.848 (95% 0.776, 0.905) ② ChatGPT (v3.5)：0.591 (95% 0.502, 0.676) ③ Google PaLM 2：0.890 (95% 0.802, 0.949) ④ Meta Llama 2：0.636 (95% 0.548, 0.718) ⑤ ChatGPT-3.5-Turbo：0.667 (95% 0.580, 0.747) ⑥ ChatGPT-4-Turbo：0.840 (95% 0.766, 0.898) ⑦ Gemini-1.0-pro：0.819 (95% 0.743, 0.881) ⑧ Llama 3：0.894 (95% 0.829, 0.941) ⑨ Claude 3 Opus：0.857 (95% 0.785, 0.912) 3.Title/Abstract（Menon 2022） (1) Sensitivity（95%CI） ① ChatGPT (v4.0)：0.932 (95% 0.847, 0.977) ② ChatGPT (v3.5)：0.315 (95% 0.211, 0.434) ③ Google PaLM 2：0.000 (95% 0.000, 0.054) ④ Meta Llama 2：0.808 (95% 0.699, 0.891) ⑤ ChatGPT-3.5-Turbo：0.470 (95% 0.352, 0.590) ⑥ ChatGPT-4-Turbo：0.760 (95% 0.646, 0.852) ⑦ Gemini-1.0-pro：0.950 (95% 0.872, 0.987) ⑧ Llama 3：0.960 (95% 0.886, 0.992) ⑨ Claude 3 Opus：0.550 (95% 0.429, 0.667) (2) Specificity（95%CI） ① ChatGPT (v4.0)：0.900 (95% 0.824, 0.951) ② ChatGPT (v3.5)：1.000 (95% 0.964, 1.000) ③ Google PaLM 2：1.000 (95% 0.958, 1.000) ④ Meta Llama 2：0.840 (95% 0.753, 0.906) ⑤ ChatGPT-3.5-Turbo：0.960 (95% 0.901, 0.989) ⑥ ChatGPT-4-Turbo：0.960 (95% 0.901, 0.989) ⑦ Gemini-1.0-pro：0.910 (95% 0.836, 0.958) ⑧ Llama 3：0.890 (95% 0.812, 0.944) ⑨ Claude 3 Opus：0.990 (95% 0.946, 1.000) (3) Accuracy（95%CI） ① ChatGPT (v4.0)：0.913 (95% 0.861, 0.951) ② ChatGPT (v3.5)：0.711 (95% 0.637, 0.777) ③ Google PaLM 2：0.569 (95% 0.486, 0.648) ④ Meta Llama 2：0.827 (95% 0.762, 0.880) ⑤ ChatGPT-3.5-Turbo：0.753 (95% 0.682, 0.816) ⑥ ChatGPT-4-Turbo：0.876 (95% 0.817, 0.921) ⑦ Gemini-1.0-pro：0.927 (95% 0.877, 0.961) ⑧ Llama 3：0.920 (95% 0.869, 0.955) ⑨ Claude 3 Opus：0.804 (95% 0.737, 0.861) | 1.Time to decision: Processes 200 abstracts in 10–20 minutes, with parallel processing capability. 2.Saves significantly on cost: (approx. $16.2 for original models, $7.1 for newer models) compared to manual screening. |
| GPT-3.5 Turbo and GPT-4 Turbo in Title and Abstract Screening for Systematic Reviews | Takehiko Oami (2025) | GPT-4T, GPT-3.5T | API | Zero-shot, criteria embedding | No | Model intergroup comparison | 1.Sensitivity: GPT-3.5T: 0.83 (95% CI 0.67 - 0.92), GPT-4T: 0.85 (95% CI 0.63 - 0.95) 2.Specificity: GPT-3.5T: 0.51 (95% CI 0.39 - 0.63) GPT-4T: 0.98 (95% CI 0.97 - 0.99) | 1.Time to decision: GPT-3.5 Turbo：0.9 min；GPT-4 Turbo：1.6 min 2.Automation level: High |
| How Well Do ChatGPT and Claude Perform in Study Selection for Systematic Review in Obstetrics | Suppachai Insuk(2025) | GPT-4o and Claude-3.5-Sonnet | Web-based | Chain-of-thought, role prompting, few-shot, criteria embedding | No | Model intergroup comparison, comparison with novice researchers without systematic review experience, and internal variable control. | 1.Title/Abstract: (1) Sensitivity (Recall) ① GPT-4o: 0.8182 (95% CI: 0.7991–0.8359) ② Claude-3.5-Sonnet: 0.8182 (95% CI: 0.7991–0.8359) (2) Specificity ① GPT-4o: 0.9116 (95% CI: 0.8966–0.9244) ② Claude-3.5-Sonnet: 0.9426 (95% CI: 0.9300–0.9528) (3) Precision ① GPT-4o: 0.1656 (95% CI: 0.1486–0.1841) ② Claude-3.5-Sonnet: 0.2411 (95% CI: 0.2213–0.2621) (4) Accuracy ① GPT-4o: 0.9138 (95% CI: 0.8994–0.9263) ② Claude-3.5-Sonnet: 0.9448 (95% CI: 0.9328–0.9547) (5) F1-Score ① GPT-4o: 0.2755 (95% CI: 0.2495–0.3016) ② Claude-3.5-Sonnet: 0.3724 (95% CI: 0.3459–0.3987) (6) Negative Predictive Value (NPV) ① GPT-4o: 0.9959 (95% CI: 0.9915–0.9980) ② Claude-3.5-Sonnet: 0.9961 (95% CI: 0.9918–0.9982) 2.Full‑Text Review (1) Sensitivity (Recall) ① GPT-4o (Short Prompt): 1.00 (95% CI: 0.8095–1.0000) ② Claude-3.5-Sonnet (Long Prompt): 1.00 (95% CI: 0.8095–1.0000) (2) Specificity ① GPT-4o (Short Prompt): 0.8333 (95% CI: 0.5520–0.9530) ② Claude-3.5-Sonnet (Long Prompt): 0.50 (95% CI: 0.2538–0.7462) (3) Precision ① GPT-4o (Short Prompt): 0.8182 (95% CI: 0.6095–0.9284) ② Claude-3.5-Sonnet (Long Prompt): 0.60 (95% CI: 0.3913–0.7778) (4) Accuracy ① GPT-4o (Short Prompt): 0.9048 (95% CI: 0.7109–0.9735) ② Claude-3.5-Sonnet (Long Prompt): 0.7143 (95% CI: 0.5004–0.8619) (5) F1-Score ① GPT-4o (Short Prompt): 0.90 (95% CI: 0.7797–0.9759) ② Claude-3.5-Sonnet (Long Prompt): 0.75 (95% CI: 0.6187–0.8480) (6) Negative Predictive Value ① GPT-4o (Short Prompt): 1.00 (95% CI: 0.8095–1.0000) ② Claude-3.5-Sonnet (Long Prompt): 1.00 (95% CI: 0.8095–1.0000) | 1. Screening burden: Title/Abstract: A total of 1,648 articles, with 29 articles proceeding to full-text review. Human involvement is required for discussing discrepancies and removing duplicates. 2. Time to decision: Title/Abstract: 5-10 seconds per record; Full-text: 15-40 seconds per article. 3. Automation level: Assisted automation |
| Is Large Language Model-Assisted Citation Screening Feasible in a Scoping Review on Nonpharmacological Interventions for Delirium in Patients With Cancer? | Yoshiyasu Ito(2025) | GPT-4T, GPT-4o, o1 | API | Zero-shot | No | Model intergroup comparison | 1. Title/Abstract (1)Sensitivity ① GPT-4T: 0.43(95%CI 0.06-0.80) ② GPT-4o: 0.71(95%CI 0.38-1.00) ③ o1: 1.00(95% CI 1.00-1.00) (2)Specificity ① GPT-4T: 0.99(95%CI 0.99-1.00) ② GPT-4o: 0.97(95%CI 0.96-0.98) ③ o1: 0.91(95% CI 0.89-0.92) (3)Precision ① GPT-4T: 0.23(95% CI 0.00-0.46) ② GPT-4o: 0.11(95% CI 0.02-0.20) ③ o1: 0.05(95% CI 0.02-0.09) 2. Full-text (1)Sensitivity ① GPT-4T: 1.00(95%CI 1.00-1.00) ② GPT-4o: 1.00(95%CI 1.00-1.00) ③ o1: 1.00(95%CI 1.00-1.00) (2)Specificity ① GPT-4T: 0.99(95%CI 0.99-1.00) ② GPT-4o: 0.97(95%CI 0.96-0.98) ③ o1: 0.90(95%CI 0.89-0.92) (3)Precision ① GPT-4T: 0.08(95%CI 0.00-0.22) ② GPT-4o: 0.02(95%CI 0.00-0.07) ③ o1: 0.01(95%CI 0.00-0.02) | 1.Workload saved: reduce the overall screening workload 2.Screening burden: increase the burden of review by introducing large volumes of false positives |
| Large language models for abstract screening in systematic- and scoping reviews: A diagnostic test accuracy study | Christian Hedeager Krag(2024) | GPT-4o, GPT-4T, GPT-3.5, Claude-3-Opus, Claude-3-Sonnet and Claude-3-Haiku | API | Zero-shot,Criteria Embedding | No | Model intergroup comparison, internal methodological control. | 1.Abstract Inclusion for Systematic Review (1)Sensitivity ① GPT-3.5: 0.42 (95%CI 95%CI 0.15-0.72) ② Claude3-Haiku: 1.0 (95%CI 95%CI 0.74-1.0) ③ Claude3-Sonnet: 0.83 (95%CI 95%CI 0.52-0.98) ④ GPT-4T: 0.42 (95%CI 95%CI 0.15-0.72) ⑤ GPT-4o: 0.33 (95%CI 95%CI 0.10-0.65) ⑥ Claude3-Opus: 0.50 (95%CI 95%CI 0.21-0.79) (2)Specificity ① GPT-3.5: 0.91 (95%CI 0.88-0.93) ② Claude3-Haiku: 0.58 (95%CI 0.53-0.62) ③ Claude3-Sonnet: 0.70 (95%CI 0.65-0.74) ④ GPT-4T: 0.99 (95%CI 0.98-1.0) ⑤ GPT-4o: 0.98 (95%CI 0.97-0.99) ⑥ Claude3-Opus: 0.98 (95%CI 0.97-0.99) (3)AUC ① GPT-3.5: 0.86 ② Claude3-Haiku: 0.82 ③ Claude3-Sonnet: 0.86 ④ GPT-4T: 0.91 ⑤ GPT-4o: 0.889 ⑥ Claude3-Opus: 0.90 2.Specific Feature of Interest Detection in Scoping Review (1)Sensitivity ① GPT-3.5: 0.80 (95%CI 0.721-0.865) ② Claude3-Haiku: 0.792 (95%CI 0.712-0.858) ③ Claude3-Sonnet: 0.838 (95%CI 0.764-0.897) ④ GPT-4T: 0.738 (95%CI 0.654-0.812) ⑤ GPT-4o: 0.777 (95%CI 0.696-0.845) ⑥ Claude3-Opus: 0.769 (95%CI 0.687-0.839) (2)Specificity ① GPT-3.5: 0.866 (95%CI 0.859-0.872) ② Claude3-Haiku: 0.928 (95%CI 0.923-0.933) ③ Claude3-Sonnet: 0.331 (95%CI 0.322-0.34) ④ GPT-4T: 0.997 (95%CI 0.996-0.998) ⑤ GPT-4o: 0.982 (95%CI 0.98-0.985) ⑥ Claude3-Opus: 0.622 (95%CI 0.613-0.631) (3)Accuracy ① GPT-3.5: 0.865 (95%CI 0.858-0.871) ② Claude3-Haiku: 0.926 (95%CI 0.921-0.931) ③ Claude3-Sonnet: 0.337 (95%CI 0.328-0.345) ④ GPT-4T: 0.994 (95%CI 0.993-0.995) ⑤ GPT-4o: 0.98 (95%CI 0.977-0.983) ⑥ Claude3-Opus: 0.623 (95%CI 0.614-0.632) | 1. Workload saved: Up to 99% of manual screening workload can be saved. The LLM pre-screening mode alone can save 55% of the workload (with no missed studies). 2. Screening burden (LLM-assisted method, GPT-4T + GPT-4o): Human reviewers only need to arbitrate 183 abstracts. 3. Time to decision: The conventional method takes 518 hours, while the LLM-assisted method reduces this to 5.2 hours. 4. Automation level:   - Using GPT-4T + GPT-4o achieves an automation rate of 91%** (with an 8% error rate).   - The LLM pre-screening mode achieves an automation rate of 55% (with a 0% error rate). 5. Cost saved: Total savings amount to approximately $20,838.5 USD. The cost of the LLM-assisted method is only 1/60th of the conventional method. |
| LitAutoScreener: Development and Validation of an Automated Literature Screening Tool in Evidence-Based Medicine Driven by Large Language Models | Yiming Tao(2025) | GPT-4o, kimi moonshot-1-128k and deepseek-chat-2.5 | API | Few-shot, chain-of-thought | No | Model intergroup comparison, internal methodological control, and workflow comparison. | 1.Title/Abstract (1) Sensitivity ① GPT (GPT-4o): 1 ② Kimi (moonshot-v1-128k): 0.9913 ③ DeepSeek (deepseek-chat-2.5): 0.9826 (2) Precision ① GPT (GPT-4o): 0.9801 ② Kimi (moonshot-v1-128k): 0.9688 ③ DeepSeek (deepseek-chat-2.5): 0.9798 (3) Accuracy ① GPT (GPT-4o): 0.9938 ② Kimi (moonshot-v1-128k): 0.9894 ③ DeepSeek (deepseek-chat-2.5): 0.9885 2.Full-text (1) Sensitivity ① GPT (GPT-4o): 1 ② Kimi (moonshot-v1-128k): 1 ③ DeepSeek (deepseek-chat-2.5): 1 (2) Precision ① GPT (GPT-4o): 1 ② Kimi (moonshot-v1-128k): 1 ③ DeepSeek (deepseek-chat-2.5): 0.9911 (3) Accuracy ① GPT (GPT-4o): 1 ② Kimi (moonshot-v1-128k): 1 ③ DeepSeek (deepseek-chat-2.5): 0.9945 | 1. Workload saved: (1) Title/abstract screening: Saves approximately 83.3%–96.7% of time. (2) Full‑text screening: Saves 80% of time. 2. Screening burden: Human reviewers only need to verify the results after automated screening. 3. Time to decision: Title/abstract: 1–5 seconds per article (compared to 30–60 seconds manually). Full‑text: 60 seconds per article (compared to 5 minutes manually). 4. Automation level:Fully automated. |
| Performance of a Large Language Model in Screening Citations | Takehiko Oami(2024) | GPT-4T | API | Zero-shot, chain-of-thought, criteria embedding, role promoting | No | Model intergroup comparison | Title/Abstract: (1) Sensitivity  ① Original prompt (primary analysis): 0.75 (95%CI 0.43-0.92) ② Modified prompt (post hoc): 0.89 (95%CI 0.74-0.95) ③ Modified prompt + Majority-vote strategy (post hoc): 0.91 (95%CI 0.77-0.97) ④ Original prompt + Chain-of-thought strategy (post hoc): 0.71 (95%CI 0.45-0.88) ⑤ Modified prompt + Chain-of-thought strategy (post hoc): 0.87 (95%CI 0.67-0.96) (2) Specificity ① Original prompt (primary analysis): 0.99 (95%CI 0.99-0.99) ② Modified prompt (post hoc): 0.98 (95%CI 0.97-0.99) ③ Modified prompt + Majority-vote strategy (post hoc): 0.98 (95%CI 0.96-0.99) ④ Original prompt + Chain-of-thought strategy (post hoc): 0.99 (95%CI 0.98-0.99) ⑤ Modified prompt + Chain-of-thought strategy (post hoc): 0.98 (95%CI 0.96-0.99) | 1.Workload saved: Approximately 92.4% of the processing time was saved. 2.Screening burden: The proportion of publications requiring manual review ranged from 0.23% to 4.7% of the total. 3.Time to decision: The LLM-assisted method required 1.3 minutes (95% CI 1.28-1.32) to process 100 studies, compared to 17.2 minutes (95% CI 14.2-18.6) for the conventional method, resulting in a mean difference of −15.25 minutes (95% CI −17.70 to −12.79). 4.Automation level: High-level automation. |
| Sensitivity, specificity and avoidable workload of using a large language models for title and abstract screening in systematic reviews and meta-analyses | Viet-Thi Tran(2023) | GPT-3.5T | API | Zero-shot | No | Internal sensitivity analysis comparison. | Title/Abstract (1) Reference standard 1 ① Sensitivity - ChatGPT： Sommer et al. (2022)：0.813 (95%CI 0.771-0.849) Sommer et al. (2023)：0.981 (95%CI 0.976-0.985) Yaacoub et al. (unpublished)：0.920 (95%CI 0.913-0.927) Kiesswetter et al. (2023)：0.992 (95%CI 0.990-0.994) Sbidian et al. (2023)：0.916 (95%CI 0.907-0.923) Pooled statistic：0.953 (95%CI 0.862-0.985) ② Specificity - ChatGPT： Sommer et al. (2022)：0.566 (95%CI 0.508-0.622) Sommer et al. (2023)：0.386 (95%CI 0.302-0.478) Yaacoub et al. (unpublished)：0.522 (95%CI 0.478-0.566) Kiesswetter et al. (2023)：0.091 (95%CI 0.064-0.127) Sbidian et al. (2023)：0.248 (95%CI 0.217-0.283) Pooled statistic：0.330 (95%CI 0.165-0.551) (2) Reference standard 2 ① Sensitivity - ChatGPT： Sommer et al. (2022)：0.875 (95%CI 0.839-0.905) Sommer et al. (2023)：0.991 (95%CI 0.987-0.993) Yaacoub et al. (unpublished)：0.946 (95%CI 0.940-0.951) Kiesswetter et al. (2023)：0.996 (95%CI 0.995-0.998) Sbidian et al. (2023)：0.925 (95%CI 0.917-0.932) Pooled statistic：0.971 (95%CI 0.896-0.992) ② Specificity - ChatGPT： Sommer et al. (2022)：0.660 (95%CI 0.603-0.712) Sommer et al. (2023)：0.386 (95%CI 0.302-0.478) Yaacoub et al. (unpublished)：0.577 (95%CI 0.535-0.622) Kiesswetter et al. (2023)：0.097 (95%CI 0.070-0.134) Sbidian et al. (2023)：0.317 (95%CI 0.283-0.354) Pooled statistic：0.377 (95%CI 0.184-0.619) | 1.Workload saved: Overall reduction of 50% – 65%, and 11% – 21% when replacing one reviewer. 2.Screening burden: Workload reduced by 50% – 65%. 3.Time to decision: Average savings of 4 – 17 hours per review (when replacing one reviewer) or 5 – 55 hours (when limiting manual screening). 4.Automation level: Zero-shot automated screening. |
| Streamlining systematic reviews with large language models using prompt engineering and retrieval augmented generation | Fouad Trad(2025) | GPT-4 | API | Zero-shot, role promoting, criteria emending | No | Comparison with Rayyan AI | 1. Title/Abstract Screening (1) Sensitivity ① Rayyan (Threshold A): 0.95 (FNR = 0.05) ② Rayyan (Threshold B): 1.00 (FNR = 0) ③ LLM (GPT-4): 1.00 (FNR = 0) 2. Full-Text Screening (Only LLM) (1) Sensitivity : 1.00 (FNR = 0) (2) Specificity: 0.996 (3) Precision: 0.256 (4) Accuracy: ~0.982 (5) F1-score: ~0.408 3. Overall Process (Title/Abstract + Full-Text, Only LLM) (1) Sensitivity: 1.00 (2) Specificity: 0.996 (3) Precision(PPV): 0.256 | 1.Workload saved: Overall workload reduced by 95.5%. 2.Screening burden: The traditional method required manual screening of titles/abstracts (14,439 articles) and full texts (1,680 articles). The Rayyan method (Threshold A) required manual screening of 3,470 titles/abstracts. After LLM screening, only 78 full-text articles required manual review. 3.Time to decision: Total time for the traditional method: 564.4 hours; Total time for the Rayyan system (Threshold A): 54.7 hours; Total time for the LLM system: 25.5 hours. 4.Automation level: The LLM system is highly automated. The Rayyan system is semi-automated and limited to the title/abstract stage. |
| Validation of large language models (Llama 3 and ChatGPT-4o mini) for title and abstract screening in biomedical systematic reviews | Adriana López-Pineda(2025) | GPT-4o mini, Llama 3 70B Instruct | API | Zero-shot, role promoting | No | Model intergroup comparison, internal methodological control. | Title/Abstract (1) Sensitivity ①GPT GPT_0: 0.562 (95%CI 0.453–0.671) GPT_1: 0.562 (95%CI 0.453–0.671) GPT_2: 0.562 (95%CI 0.453–0.671) ②Llama 3 LLA_0: 0.750 (95%CI 0.655–0.845) LLA_1: 0.750 (95%CI 0.655–0.845) LLA_2: 0.775 (95%CI 0.683–0.867) (2) Specificity ①GPT GPT_0: 0.950 (95%CI 0.936–0.964) GPT_1: 0.948 (95%CI 0.934–0.962) GPT_2: 0.951 (95%CI 0.937–0.965) ②Llama 3 LLA_0: 0.914 (95%CI 0.896–0.932) LLA_1: 0.910 (95%CI 0.891–0.929) LLA_2: 0.914 (95%CI 0.896–0.932) (3) Precision ①GPT GPT_0: 0.500 (95%CI 0.397–0.603) GPT_1: 0.489 (95%CI 0.387–0.591) GPT_2: 0.506 (95%CI 0.402–0.610) ②Llama 3 LLA_0: 0.435 (95%CI 0.352–0.518) LLA_1: 0.426 (95%CI 0.344–0.508) LLA_2: 0.443 (95%CI 0.361–0.525) (4) Accuracy ①GPT GPT_0: 0.919 GPT_1: 0.917 GPT_2: 0.920 ②Llama 3 LLA_0: 0.900 LLA_1: 0.897 LLA_2: 0.902 (5) F1-score ①GPT GPT_0: 0.529 GPT_1: 0.519 GPT_2: 0.533 ②Llama 3 LLA_0: 0.548 LLA_1: 0.543 LLA_2: 0.564 | 1.Workload saved: Reduced by 30%–70% 2.Screening burden: Decreased from 1,081 records to approximately 162 records 3.Automation level: Semi-automated |

LLM: Large language model; API: Application programming interface; BERT: Bidirectional encoder representations from transformers; GPT-4T: GPT-4 Turbo; GPT-4o: GPT-4 Omni.
