## Supplementary material for "Performance of Large Language Models in Automated Medical Literature Screening: A Systematic Review and Meta-analysis": Table 3

Table3. The literature included in the quality assessment was conducted using the PROBAST + AI tool.

| **Title** | **Author(year)** | **Development** | | | | | | | **Evaluation** | | | | | | | **Whole** | | | |
| --- | --- | --- | --- | --- | --- | --- | --- | --- | --- | --- | --- | --- | --- | --- | --- | --- | --- | --- | --- |
|  |  | Participants &Data Source | | Predictors | | Outcome | | Analysis | Participants &Data Source | | Predictors | | Outcome | | Analysis | Development Section | | Evaluation section | |
|  |  | A | B | A | B | A | B | A | C | B | C | B | C | B | C | Quality | applicability | Bias risk | applicability |
| Accuracy of Large Language Models for Literature Screening in Thoracic Surgery: Diagnostic Study | Zhang-Yi Dai(2025) | + | √ | + | √ | + | √ | + | + | √ | + | √ | + | √ | + | + | + | + | + |
| Assessing the performance of large language models in literature screening for pharmacovigilance a comparative study | Dan Li (2024) | + | √ | + | √ | + | √ | + | + | √ | + | √ | + | √ | + | + | + | + | + |
| Automated Literature Screening for Hepatocellular Carcinoma Treatment Through Integration of 3 Large Language Models：Methodological Study | Chen Pan (2025) | + | √ | + | √ | + | √ | + | + | √ | + | √ | + | √ | + | + | + | + | + |
| Automated Paper Screening for Clinical Reviews Using Large Language Models：Data Analysis Study | Eddie Guo (2024) | + | √ | - | √ | - | √ | + | + | √ | - | √ | - | √ | + | - | + | - | + |
| Can large language models replace humans in systematic reviews？Evaluating GPT-4's efficacy in screening and extracting data from peer-reviewed and grey literature in multiple languages | QusaiKhraisha (2023) | + | √ | + | √ | + | √ | - | + | √ | + | √ | + | √ | + | - | + | + | + |
| Development and evaluation of prompts for a large language model to screen titles and abstracts in a living systematic review | Ava Homiar (2025) | + | √ | + | √ | + | √ | + | + | √ | + | √ | + | √ | + | + | + | + | + |
| Enhancing AI for citation screening in literature reviews：Improving accuracy with ensemble models | Zhihong Zhang (2025) | + | √ | + | √ | + | √ | + | + | √ | + | √ | + | √ | + | + | + | + | + |
| Evaluating GPT Models for Automated Literature Screening in Wastewater-Based Epidemiology | KasebaChibwe (2024) | + | √ | + | √ | + | √ | + | + | √ | + | √ | + | √ | - | + | + | + | + |
| Evaluating the efectiveness of large language models in abstract screening: a comparative analysis | Michael Li (2024) | + | √ | + | √ | + | √ | + | + | √ | + | √ | + | √ | + | + | + | + | + |
| GPT-3.5 Turbo and GPT-4 Turbo in Title and Abstract Screening for Systematic Reviews | Takehiko Oami (2025) | + | √ | - | √ | + | √ | ？ | + | √ | - | √ | + | √ | - | - | + | - | + |
| How Well Do ChatGPT and Claude Perform in Study Selection for Systematic Review in Obstetrics | Suppachai Insuk (2025) | + | √ | + | √ | + | √ | + | + | √ | + | √ | + | √ | - | + | + | - | + |
| Is Large Language Model-Assisted Citation Screening Feasible in a Scoping Review on Nonpharmacological Interventions for Delirium in Patients With Cancer? | Yoshiyasu Ito (2025) | + | √ | - | √ | + | √ | ？ | + | √ | - | √ | + | √ | ？ | - | + | - | + |
| Large language models for abstract screening in systematic- and scoping reviews: A diagnostic test accuracy study | Christian Hedeager Krag (2024) | + | √ | + | √ | + | √ | + | + | √ | + | √ | + | √ | + | + | + | + | + |
| LitAutoScreener: Development and Validation of an Automated Literature Screening Tool in Evidence-Based Medicine Driven by Large Language Models | Yiming Tao (2025) | + | √ | + | √ | + | √ | + | + | √ | + | √ | + | √ | + | + | + | + | + |
| Performance of a Large Language Model in Screening Citations | Takehiko Oami (2024) | + | √ | + | √ | + | √ | ？ | + | √ | + | √ | + | √ | - | ？ | + | - | + |
| Sensitivity, specificity and avoidable workload of using a large language models for title and abstract screening in systematic reviews and meta-analyses | Viet-Thi Tran (2023) | + | √ | + | √ | + | √ | + | + | √ | + | √ | + | √ | + | + | + | + | + |
| Streamlining systematic reviews with large language models using prompt engineering and retrieval augmented generation | Fouad Trad (2025) | + | √ | + | √ | + | √ | - | + | √ | + | √ | + | √ | + | - | + | + | + |
| Validation of large language models (Llama 3 and ChatGPT-4o mini) for title and abstract screening in biomedical systematic reviews | Adriana López-Pineda (2025) | + | √ | + | √ | + | √ | ？ | + | √ | + | √ | + | √ | - | ？ | + | - | + |

A: Quality assessment; B: Evaluation of suitability; C: Bias risk assessment; +:High quality/low risk of bias; -: low quality/high risk of bias; ?:nuclear; √：High versatility; ×：low versatility.
