## Supplementary material for "Performance of Large Language Models in Automated Medical Literature Screening: A Systematic Review and Meta-analysis": Table 4

Table 4. The literature included in the quality assessment was conducted using the QUADAS 2 tool.

|  | **Risk of Bias** | | | | **Applicability Concerns** | | | |
| --- | --- | --- | --- | --- | --- | --- | --- | --- |
| **Title** | **Patient selection** | **Index test** | **Reference standard** | **Flow and timing** | **Patient selection** | **Index test** | **Reference standard** | **Flow and timing** |
| Is Large Language Model-Assisted Citation Screening Feasible in a Scoping Review on Nonpharmacological Interventions for Delirium in Patients With Cancer? | Low | Low | Low | Low | Low | Low | Low | - |
| Large language models for abstract screening in systematic- and scoping reviews: A diagnostic test accuracy study | Low | Low | Low | Low | Low | Low | Low | - |
| Evaluating the efectiveness of large language models in abstract screening: a comparative analysis | Unclear | Unclear | Low | Low | Unclear | Low | Low | - |
| Development and evaluation of prompts for a large language model to screen titles and abstracts in a living systematic review | Low | Low | Low | Low | Low | Low | Low | - |
| Sensitivity, specificity and avoidable workload of using a large language models for title and abstract screening in systematic reviews and meta-analyses | Low | Low | Low | Low | Low | Low | Low | - |
| Accuracy of Large Language Models for Literature Screening in Thoracic Surgery: Diagnostic Study | Unclear | Unclear | Unclear | Unclear | Unclear | Low | Low | - |
| Automated Literature Screening for Hepatocellular Carcinoma Treatment Through Integration of 3 Large Language Models: Methodological Study | Low | Low | Low | Low | Low | Low | Low | - |
| LitAutoScreener: Development and Validation of an Automated Literature Screening Tool in Evidence-Based Medicine Driven by Large Language Models | Unclear | Unclear | Unclear | Unclear | Unclear | Low | Low | - |
| Automated Paper Screening for Clinical Reviews Using Large Language Models: Data Analysis Study | Low | Low | Low | Low | Low | Low | Low | - |
| Assessing the performance of large language models in literature screening for pharmacovigilance: a comparative study | Unclear | Unclear | Unclear | Unclear | High | Low | Low | - |
| Enhancing AI for citation screening in literature reviews: Improving accuracy with ensemble models | Unclear | Unclear | Unclear | Unclear | Unclear | Low | Low | - |
| Evaluating GPT Models for Automated Literature Screening in Wastewater-Based Epidemiology | Low | Low | Low | Low | Low | Low | Low | - |
| GPT-3.5 Turbo and GPT-4 Turbo in Title and Abstract Screening for Systematic Reviews | Unclear | Unclear | Unclear | Unclear | Unclear | Low | Low | - |
| Validation of large language models (Llama 3 and ChatGPT-4o mini) for title and abstract screening in biomedical systematic reviews | Low | Low | Low | Low | Low | Low | Low | - |
| Performance of a Large Language Model in Screening Citations | Low | Low | Low | Low | Low | Low | Low | - |
| Can large language models replace humans in systematic reviews? Evaluating GPT-4's efficacy in screening and extracting data from peer-reviewed and grey literature in multiple languages | Unclear | Unclear | Unclear | Unclear | Unclear | Low | Low | - |
| How Well Do ChatGPT and Claude Perform in Study Selection for Systematic Review in Obstetrics | Low | Low | Low | Low | Low | Low | Low | - |
| Streamlining systematic reviews with large language models using prompt engineering and retrieval augmented generation | Low | Low | Low | Low | Low | Low | Low | - |
